# Copy-number variants as modulators of common disease susceptibility

**DOI:** 10.1101/2023.07.31.23293408

**Authors:** Chiara Auwerx, Maarja Jõeloo, Marie C. Sadler, Nicolò Tesio, Sven Ojavee, Charlie J. Clark, Reedik Mägi, Estonian Biobank Research Team, Alexandre Reymond, Zoltán Kutalik

## Abstract

**Background:** Copy-number variations (CNVs) have been associated with rare and debilitating genomic syndromes but their impact on health later in life in the general population remains poorly described.

**Methods:** Assessing four modes of CNV action, we performed genome-wide association scans (GWASs) between the copy-number of CNV-proxy probes and 60 curated ICD-10 based clinical diagnoses in 331,522 unrelated white UK Biobank participants with replication in the Estonian Biobank.

**Results:** We identified 73 signals involving 40 diseases, all of which indicating that CNVs increased disease risk and caused earlier onset. Even after correcting for these signals, a higher CNV burden increased risk for 18 disorders, mainly through the number of deleted genes, suggesting a polygenic CNV architecture. Number and identity of genes disturbed by CNVs affected their pathogenicity, with many associations being supported by colocalization with both common and rare single nucleotide variant association signals. Dissection of association signals provided insights into the epidemiology of known gene-disease pairs (e.g., deletions in *BRCA1* and *LDLR* increased risk for ovarian cancer and ischemic heart disease, respectively), clarified dosage mechanisms of action (e.g., both increased and decreased dosage of 17q12 impacts renal health), and identified putative causal genes (e.g., *ABCC6* for kidney stones). Characterization of the pleiotropic pathological consequences of recurrent CNVs at 15q13, 16p13.11, 16p12.2, and 22q11.2 in adulthood indicated variable expressivity of these regions and the involvement of multiple genes.

**Conclusions:** Our results shed light on the prominent role of CNVs in determining common disease susceptibility within the general population and provide actionable insights allowing to anticipate later-onset comorbidities in carriers of recurrent CNVs.

## BACKGROUND

Copy-number variants (CNVs) refer to duplicated or deleted DNA fragments (≥ 50bp) and represent an important source of inter-individual variation [1,2]. As a highly diverse mutational class, they can alter the copy-number of dosage sensitive genes, induce gain- or loss-of-function (LoF) through gene fusion or truncation, unmask recessive alleles, or disrupt regulatory sequences, thereby representing potent phenotypic modifiers [3]. As such, their role in human disease has mainly been studied in clinically ascertained cohorts often presenting with congenital anomalies and/or severe neurological (e.g., developmental delay and intellectual disability, epilepsy) or psychiatric (e.g., autism or schizophrenia) symptoms [4–7] and today, close to 100 genomic disorders (i.e., disease caused by genomic rearrangements) have been described [8,9]. Despite their deleteriousness, some of these CNVs flanked by repeats recurrently appear and remain at a low but stable frequency in the population [10].

The emergence of large biobanks coupling genotype to phenotype data has fostered the study of CNVs in the general population. Whole genome sequencing represents the best approach to characterize the full human CNV landscape [1,11,12] but current long- and short-read sequencing association studies have limited samples size [13–15]. Alternatively, larger sample sizes are available for exome sequencing data, offering the possibility to assess the phenotypic consequence of small CNVs [16,17], while microarray-based CNV calls are better-suited for the study of large CNVs and have been successfully used in association studies [8,18–28]. Performing a CNV genome-wide association study (GWAS) on 57 medically relevant continuous traits in the UK Biobank (UKBB) [29], we previously identified 131 independent associations, including allelic series wherein carriers of CNVs at loci previously associated with rare Mendelian disorders exhibited subtle changes in disease-associated phenotypes but lacked the corresponding clinical diagnosis [23]. Paralleling findings for point mutations [30–33], this supports a model of variable expressivity, where CNVs can cause a wide spectrum of phenotypic alteration ranging from severe, early-onset diseases to mild subclinical symptoms, opening the question as to whether these loci are also associated with common diseases.

Unlike continuous traits that can be objectively measured in all participants, population cohorts, such as UKBB, have low numbers of diseased individuals [34]. Moreover, defining cases relies on the arbitrary dichotomization of complex underlying pathophysiological processes [35]. Beyond the inherent loss of power associated to usage of binary variables [36], cases might be missed because an individual did not consult a physician, was misdiagnosed due to atypical clinical presentation, or is in a prodromal disease phase. Studies investigating CNV-disease associations in the general population have either focused on only few diseases [27,37] or well-established recurrent CNVs [20,38,39]. Alternatively, high-throughput studies have assessed a broad range of continuous and binary traits simultaneously [17,24,25] without any precautions to accommodate the aforementioned challenges. To date, the largest disease CNV-GWAS meta-analyzed ∼1,000,000 individuals [8]. While boosting power through increased sample size, it comes at the cost of extensive data harmonization, resulting in the exclusion of smaller CNVs (≤ 100kb) and broader disease categories (e.g., “immune abnormality”). Moreover, as this study includes several clinical cohorts, phenotypes are biased towards neuropsychiatric disorders (24/54 phenotypes) for which the role of CNVs is well-established [4–7].

Using tailored CNV-GWAS models mimicking four mechanisms of CNV action and time-to-event analysis, we investigate the relationship between CNVs and 60 carefully defined common diseases affecting a broad range of physiological systems in 331,522 unrelated white UKBB participants. Extensively validating our results, we report associations according to confidence tiers and take advantage of rich individual-level phenotypic data to demonstrate the contribution of CNVs to the common disease burden in the general population.

## METHODS

### 1. Study material

#### Discovery cohort: UK Biobank

The UK Biobank (UKBB) is composed of ∼500,000 volunteers (54% females) from the general UK population for which microarray-based genotyping and extensive phenotyping data – including hospital based International Classification of Diseases, 10th Revision (ICD-10) codes (up to September 2021) and self-reported conditions – are available [29]. Participants signed a broad informed consent form and data were accessed through application #16389.

#### Replication cohort: Estonian Biobank

The Estonian Biobank (EstBB) is a population-based cohort of ∼208,000 Estonian individuals (65% females; data freeze 2022v01 [12/04/2022]) for which microarray-based genotyping data and ICD-10 codes from crosslinking with national and hospital databases (up to end 2021) are available [40]. The activities of the EstBB are regulated by the Human Genes Research Act, which was adopted in 2000 specifically for the operations of the EstBB. Individual level data analysis in the EstBB was carried out under ethical approval 1.1-12/624 from the Estonian Committee on Bioethics and Human Research (Estonian Ministry of Social Affairs), using data according to release application 3-10/GI/34668 [20/12/2022] from the EstBB. All participants signed a broad informed consent form.

#### Other public resources

The PheCode Map 1.2 (beta) (https://phewascatalog.org/phecodes_icd10) was used for ICD-10 code classification [41]. Genomic regions were annotated with the NHGRI-EBI GWAS Catalog (https://www.ebi.ac.uk/gwas/; 24/11/2022) [42], and the Online Mendelian Inheritance in Man (OMIM; https://www.omim.org/; 27/07/2022) [43]. Recurrent CNV coordinates were retrieved from DECIPHER (https://www.deciphergenomics.org/) [9]. Unless specified otherwise, Neale UK Biobank single-nucleotide polymorphism (SNP)-GWASs summary statistics were used (http://www.nealelab.is/uk-biobank). Allele frequencies and genomic constraint scores (<u>p</u>robability of <u>L</u>oF Intolerance (pLI); <u>L</u>oF <u>O</u>bserved over <u>E</u>xpected <u>U</u>pper bound <u>F</u>raction (LOEUF)) originate from the Genome Aggregation Database (GnomAD; https://gnomad.broadinstitute.org/) [44]; pHaplo and pTriplo scores from [8]. Tissue-specific gene expression was assessed in the Genotype-Tissue Expression project (GTEx; https://gtexportal.org/home/) [45].

#### Software versions

CNVs were called with PennCNV v1.0.5 [46] using PennCNV-Affy (27/08/2009) and filtered based on a quality scoring pipeline [47]. Genetic analyses were conducted with PLINK v1.9 and v2.0 [48]. ANNOVAR (24/10/2019) was used to map genes to genetic regions [49]. The UCSC Genome Browser was used to determine the human genome size (GRCh37/hg19) and the LiftOver tool was used to lift over genomic coordinates [50]. Statistical analyses were performed with R v3.6.1 and graphs were generated with R v4.1.3.

### 2. CNV association studies in the UK Biobank

#### Microarray-based CNV calling

UKBB genotype microarray data were acquired from two arrays with 95% probe overlap (Applied Biosystems UK Biobank Axiom Array: 438,427 samples; Applied Biosystems UK BiLEVE Axiom Array by Affymetrix: 49,950 samples) [29] and used to call CNVs as previously described [23]. Briefly, CNVs were called using standard PennCNV settings and samples on genotyping plates with a mean CNV count per sample > 100 and samples with > 200 CNVs or single CNV > 10 Mb were excluded. Remaining CNVs were attributed a probabilistic quality score (QS) ranging from -1 (likely deletion) to 1 (likely duplication) [47]. High confidence CNVs, stringently defined by |QS| > 0.5, were retained and encoded in chromosome-wide probe-by-sample matrices (i.e., entries of 1, -1, or 0 indicate probes overlapping high confidence duplication, deletion, or no/low quality CNV, respectively) [47], which were converted into three PLINK binary file sets to accommodate association analysis according to four modes of CNV action. Details about the CNV encoding and handling of chromosome X are provided in Supplemental Note 1. Probe-level CNV frequency was calculated [23]. All results in this study are based on the human genome reference build GRCh37/hg19.

#### Case-control definition and age-at-disease onset calculation

A pool of 331,522 unrelated white British UKBB participants (54% females) was considered after excluding retracted (up to August 2020), as well as related, high missingness and non-white British samples (*used.in.pca.calculation* = 0 and *in.white.British.ancestry.subset* = 0 in Sample-QC v2 file). CNV outliers (*Microarray-based CNV calling*) and individuals reporting blood malignancies (i.e., possibly harboring somatic CNVs; UKBB field #20001: 10047, 1048, 1050, 1051, 1052, 1053, 1055, 1056, 1056; #41270: ICD-10 codes mapping to PheCode exclusion range “*cancer of lymphatic and hematopoietic tissue*”), were further excluded.

Cases and controls were assigned for 60 ICD-10-based clinical diagnoses using *diagnosis – ICD10* (#41270), *cancer code, self-reported* (#20001), and *non-cancer illness code, self-reported* (#20002) to build exclusion and inclusion lists. For each disease, we first defined all 331,522 individuals as controls. We excluded individuals with a self-reported or hospital diagnosis of a broad set of conditions that include the disease of interest, as well as related disorders/ICD-10 codes (e.g., other cancers or radio-/chemotherapy for breast cancer; mood or personality disorders for schizophrenia). We then re-introduce as cases individuals having received a restricted range of ICD-10 codes matching our disease definition. For second level ICD-10 codes, all subcodes are considered, otherwise only the specified ones. The disease burden was calculated as the number of diagnoses (out of the 60 assessed) an individual has received. For male- (prostate cancer) and female- (menstruation disorders, endometriosis, breast cancer, ovarian cancer) specific diseases, downstream analyses were conducted excluding individuals from the opposite sex.

Based on the *date at first in-patient diagnosis – ICD10* (#41280) and the individual’s *month* (#52) and *year* (#34) *of birth* (birthday assumed on average to be the 15^th^), the age at diagnosis was calculated by subtracting the earliest diagnosis date for codes on the inclusion list from the birth date and converting it to years by dividing by 365.25 to account for leap years.

#### Probe and covariate selection

Relevant covariates and probes were pre-selected to fit tailored main CNV-GWAS models and reduce computation time. For each disease, a logistic regression was fitted to explain disease probability as a function of age (#21003), sex, genotyping array, and the 40 first principal components (PCs). Nominally significantly associated covariates (p ≤ 0.05) were retained for the main analysis. CNV-proxy probes with a CNV frequency ≥ 0.01% were pruned at r^2^ > 0.9999 in PLINK_CNV_ (`–-indep-pairwise 500 250 0.9999` PLINK v2.0) to group probes at the core of CNV regions while retaining resolution at breakpoints (BPs), resulting in 18,725 probes. For each disease, 2-by-3 genotypic Fisher tests assessed dependence between disease status and probe copy-number (rows: control versus case; columns: deletion versus copy-neutral versus duplication; `–-model fisher` PLINK v1.9; TEST column `GENO`). Probes with p ≤ 0.001 and a minimum of two disease cases among CNV, duplication, or deletion carriers were retained for assessment through the mirror/U-shaped, duplication-only, or deletion-only model, respectively.

#### Genome-wide significance threshold

Due to the recurrent nature of CNVs, the 18,725 probes retained after frequency filter and pruning remain highly correlated and are thus not independent. Accounting for all of them would result in an overly strict multiple testing correction. Using an established protocol [22,23,51], we estimated the number of effective tests performed to N_eff_ = 6,633, setting the genome-wide (GW) threshold for significance at p ≤ 0.05/6,633 = 7.5 x 10^-6^. This threshold is of the same order of magnitude as what others have estimated for disease CNV-GWAS [8].

#### Main CNV-GWAS model

Association between disease risk and copy-number of CNV-proxy probes was assessed through logistic regression with Firth fallback (`--covar-variance-standardize –-glm firth-fallback omit-ref no-x-sex hide-covar -–ci 0.95` PLINK v2.0), using disease- and model-specific probes and covariates (*Probe and covariate selection*). Four association models were assessed: the mirror model assesses the additive effect of each additional copy (PLINK_CNV_); the U-shape model assesses a consistent effect of any deviation from the copy neutral state (PLINK_CNV_, using the `hetonly` option in `glm` PLINK v2.0); the duplication-only model (PLINK_DUP_) assesses the impact of a duplication while disregarding deletions; the deletion-only model (PLINK_DEL_) assesses the impact of a deletion while disregarding duplications. Odds ratios (OR) and their 95% confidence interval (CI) were harmonized (A1 to “T”; Supplemental Note 1 – Table 1), i.e., 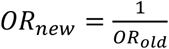 and 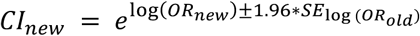, respectively. GW-significant associations (p ≤ 7.5 x 10^-6^; *Genome-wide significance threshold*) were pruned at r^2^ > 0.8 (`--indep-pairwise 3000 500 0.8` PLINK v2.0), giving priority to probes with the strongest association signal by inputting a scaled negative logarithm of association p-value as frequency (`–-read-freq` PLINK v2.0). For the U-shape model, pruning was performed using custom code by extracting probes from PLINK_CNV_ and recoding them to match U-shape numerical encoding. Number of independent signals per disease was determined by stepwise conditional analysis. Briefly, for each disease and association model, the numerical CNV genotype of the lead probe was included along selected covariates in the logistic regression model. This process was repeated until no more GW-significant signal remained.

Due to its continuous nature, the disease burden CNV-GWAS was based on linear regressions between copy-number of selected probes and the disease burden (`–-glm omit-ref no-x-sex hide-covar allow-covars` PLINK v2.0), correcting for selected covariates. Post-GWAS processing was performed as previously described [23].

#### CNV region definition and annotation

CNV region (CNVR) boundaries were defined by the most distant probe within ± 3Mb and r^2^ ≥ 0.5 of independent lead probes (`–-show-tags --tag-kb 3000 --tag-r2 0.5` PLINK v1.9; U-shape model: custom code, as described previously for pruning). Signals from different models were merged when overlapping (≥ 1bp) and involving the same disease, with CNVR boundaries defined as the maximal CNVR. Characteristics of the most significant model (i.e., “best model”) are reported. The “main model” indicates which CNV type mainly drives the association, i.e., when associations were found through multiple models, priority was given to either the duplication-only or deletion-only models, otherwise to the model yielding the lowest p-value. CNVRs were annotated with hg19 HGNC and ENSEMBL gene names using `annotate_variation.pl` from ANNOVAR (`--geneanno`). Number of genes mapped to a CNVR was calculated and set to zero for CNVRs with `REGION` not equaling “exonic”. Resulting count was used as a predictor for pleiotropy (i.e., number of associations) through linear regression.

#### Statistical confidence tiers

Following primary assessment through logistic regression (*Main CNV-GWAS model*), three statistical approaches were implemented to gauge robustness of the lead probe’s association signal. First, we assessed *post hoc* the p-value of 2-by-3 genotypic Fisher tests (*Probe and covariate selection*). Second, we transformed the binary disease status into a continuous variable by computing the response residuals of the logistic regression of disease status on disease-relevant covariates. This allowed the usage of linear regressions to estimate the effect of the CNV genotype (encoded according to all significantly associated models in the primary analysis) on disease risk. The model generating the lowest p-value for the CNV encoding is reported. Third, time-to-event analysis was used to assess whether CNVs influence age-at-disease onset. Age at last healthy measurement was calculated as age-at-disease onset for cases and date of last recorded diagnosis (30/09/2021) minus birth date converted to years for controls (*Case-control definition and age-at-disease onset calculation*). Cox proportional-hazards (CoxPH) models were fitted including disease-relevant covariates and numerically encoded CNV genotype for either of the four association models as predictors, using `coxph()` function from the R `survival` package [52]. The model with the lowest CNV genotype p-value is reported. CNV-disease associations were classified in confidence tiers depending on whether they were confirmed by 3 (tier 1), 2 (tier 2), or 1 (tier 3) of the above-described approaches at the arbitrary validation significance threshold of p ≤ 1 x 10^-4^. Validation approaches not being suited for continuous variables – which do not suffer from the same caveats as binary traits – all disease burden associations were classified as tier 1.

#### Literature-based supporting evidence

Using three literature-based approaches, we examined whether disease-associated CNVRs had previously been linked to relevant phenotypes. First, we investigated the colocalization of autosomal CNVRs with SNP-GWAS signals. GRCh38/hg38 lifted CNVR coordinates were inputted in the GWAS Catalog and associations (p ≤ 1×10^-7^) relevant to the investigated disease (i.e., synonym, continuous proxy, or major risk factor) were identified through manual curation. Second, we overlapped OMIM morbid genes (i.e., linked to an OMIM disorder; `morbidmap.txt`) with disease-associated CNVRs. Through manual curation, we flagged OMIM genes associated to Mendelian disorders sharing clinical features with the common disease associated through CNV-GWAS. Third, we examined if implicated CNVRs overlapped regions at which CNVs were found to modulate continuous traits [23] or disease risk [20,25].

### 3. Replication in the Estonian Biobank

#### CNV calling and sample selection

Autosomal CNVs were called from Illumina Global Screening Array (GSA) genotype data for 193,844 individuals that survived general quality control and had matching genotype-phenotype identifiers, matching inferred versus reported sex, a SNP-call rate ≥ 98%, and were included in the EstBB SNP imputation pipeline. CNV outliers and individuals with a reported blood malignancy were excluded, as previously described. High confidence CNV calls (|QS| > 0.5) of the 156,254 remaining individuals were encoded into three PLINK binary file sets, following the procedure described for the UKBB (CNV association studies in the UK Biobank).

#### EstBB disease definition

Disease cases and disease burden were defined similarly than in the UKBB. To account for differences in recording practices between the countries, Z12 (routine preventive screens for cancer), and D22-23 (benign skin lesions) subcodes, were removed from the exclusion list of cancer traits as they were much more frequent than in the UKBB and strongly reduced the number of controls. Due to lack of matching data in the EstBB, no self-reported diseases and cancers were used as an exclusion criterion for disease definition.

#### EstBB replication analysis

Related individuals with available CNV calls were pruned (KING kinship coefficient > 0.0884), prioritizing individuals whose disease status was least often missing, leaving 90,211 unrelated samples for the replication study. Disease-relevant covariates were selected among sex, year of birth, genotyping batch (1-11), and PC1-20. For each of the 73 UKBB signals, probes overlapping the CNVR and with an EstBB CNV, duplication, or deletion frequency ≥ 0.01%, were retained, depending on whether the mirror/U-shape, duplication-only, or deletion-only was the best UKBB model, respectively. Association studies were performed on remaining probes using disease-specific covariates and the best UKBB model, following the previously described procedure. Forty signals (55%) could not be assessed due to null/low CNV frequency, failure of the regression to converge, absence of at least one case CNV carrier, or because not mapping on the autosomes. For the remaining 33 signals, summary statistics of the probe showing the strongest association within the CNVR were retained and p-values were adjusted to account for directional concordance with UKBB effects by rewarding and penalizing signals with matching and non-matching effect size signs, respectively. Specifically, one-sided p-values were obtained as 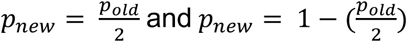 for 24 concordant and 9 non-concordant signals, respectively. Accounting for 33 testable signals, the replication threshold for significance was set at p ≤ 0.05/33 = 1.5 x 10^-3^. One-sided binomial tests (`binom.test()`) were used to assess enrichment of observed versus expected significant associations at various thresholds (*α* = 0.1 to 0.005 by steps of 0.005), with the R function arguments: *x* the number of observed signals at *α*, *n* the number of testable signals (i.e., 33), and *p* the expected probability of signals meeting *α* (i.e., *α*).

### 4. CNV region constraint analysis

Evolutionary constraint of genes overlapping disease-associated CNVRs, i.e., “disease genes” (*CNV region definition and annotation*), was assessed by comparing their pLI, LOEUF, pHaplo, and pTriplo scores to the ones of “background genes”. The latter were identified by annotating ranges of one or multiple consecutive probes with CNV frequency ≥ 0.01% with ANNOVAR (hg19 HGNC gene names) and excluding disease genes. For pLi and LOEUF, all disease genes were considered together. For pHaplo and pTriplo, two disease gene groups were considered: genes overlapping CNVRs with at least one association through the duplication-only model and genes overlapping CNVRs with at least one association through the deletion-only model. As many CNVRs associated through both models, the analysis was repeated considering genes overlapping CNVRs with at least one association through the duplication-only and none through the deletion-only model and vice-versa. Comparison with background genes was done through two-sided Wilcoxon rank-sum test.

### 5. Extended phenotypic assessment

To elaborate on specific associations, we made use of the rich phenotypic data available for UKBB participants, as detailed in Supplemental Note 2. For fine-mapping of association signals, CNV carriers were divided in subgroups based on visual inspection of CNV breakpoints and segmental duplications, as detailed in Supplemental Note 3.

#### CNV versus copy-neutral comparisons

Comparisons between groups of CNV carriers and copy-neutral individuals always exclude low quality CNV (|QS| ≤ 0.5) carriers altogether. For diseases, prevalence is estimated as 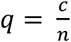, with *c* and *n* are the number of cases and total number of individuals in a group, and 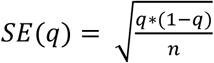. Differences in prevalence compared to copy-neutral individuals were assessed by two-sided Fisher test. For continuous traits, comparisons are based on two-sided t-tests.

### 6. CNV burden analyses

#### CNV burden association studies

In the UKBB, individual-level CNV, duplication, and deletion burden were calculated as the number of Mb or genes affected by high-confidence (|QS| > 0.5) autosomal CNVs, duplications, and deletions, respectively, as previously described [23]. Association between burden values and the 60 diseases (logistic regression) or the disease burden (linear regression), was assessed including disease-relevant covariates in the model. Accounting for the 61 evaluated traits, significance was defined at p ≤ 0.05/61 = 8.2 x 10^-4^. We next corrected burden values for CNV-GWAS signals. For each disease, CNVs, duplications, and deletions overlapping (≥ 1bp) a CNVR significantly associated with the disease of interest through CNV-GWAS were omitted from the CNV, duplication, and deletion burden calculations if the CNVR had been found to associate with the disease through the mirror/U-shape, duplication-only, or deletion-only model, respectively. Association studies were repeated using corrected burden values. Only the most significant burden types are reported in the text.

#### Relative importance of protein coding regions in mediating the burden’s effect

The average genome-wide gene density (*GD_GW_*) was estimated to 8.4 genes/Mb based on 26,289 genes (i.e., unique HGNC gene names in hg19 RefSeq, excluding microRNAs but including genes of uncertain function (i.e., “LOC”)) and a human genome length of 3,137,161,264 bp. If CNVs affecting the coding and non-coding DNA have similar effects, we expect that the impact of 1Mb affected by CNVs to be equivalent to 8.4 genes being affected by CNVs. Hence, the association effect size of the CNV burden measured in Mb (*β_Mb_*) is expected to be 8.4-times larger than the one measured in number of affected genes (*β_gene_*), i.e., 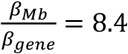. This hypothesis was tested independently for the deletion and duplication burdens for traits with at least one significant uncorrected burden association (i.e., 20 diseases + disease burden). Significant deviations from the expected ratio were assessed by t-statistic:

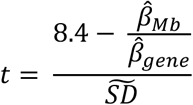

where *β̂*_*Mb*_ and *β̂*_*gene*_ are the estimated effects of the burden measured in Mb or number of genes impacted by CNVs, respectively, on the assessed trait. *SD* is the empirically observed standard deviation of the 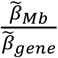 ratio, estimated based on 10,000 simulations of 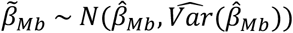 and 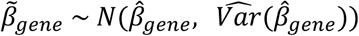. P-values were computed based on a two-sided one sample t-test and deemed significant at p ≤ 0.05/21 = 2.4 x 10^-3^. This analysis was repeated for the modified burden definitions not accounting for CNVs overlapping disease-associated CNVRs.

## RESULTS

### The spectrum of common diseases in the UK Biobank

To mitigate issues related to disease definition, we used a three-step approach to designate cases and controls in the UKBB (Figure 1A; top). Starting from 331,522 unrelated white British individuals, we defined cases based on a narrow list of hospital-based diagnoses (i.e., ICD-10 codes) and excluded self-reported cases, as well as self-reported and hospital diagnoses of related conditions. Sixty disorders spanning 12 ICD-10 chapters were selected to cover a wide range of physiological systems, favoring conditions with sufficiently large sample size and a likely genetic basis (Figure 1B; Figure S1A; Table S1). Except for systemic lupus erythematosus (N = 422) and polycystic kidney disease (N = 454), all diseases had over 500 cases. Nineteen diseases had a case count > 10,000, with osteoarthrosis (N = 62,175) and essential hypertension (N = 97,860) being the most frequent. Seven diseases had a median age of onset ≤ 60 years, predominantly female reproductive disorders, autoimmune conditions, and psychiatric disorders. Conversely, the nine diseases with a median age at onset ≥ 70 years were mainly degenerative disorders of the brain, eye, and kidney, overall aligning align with epidemiological knowledge of the respective diseases.

**Figure 1.**
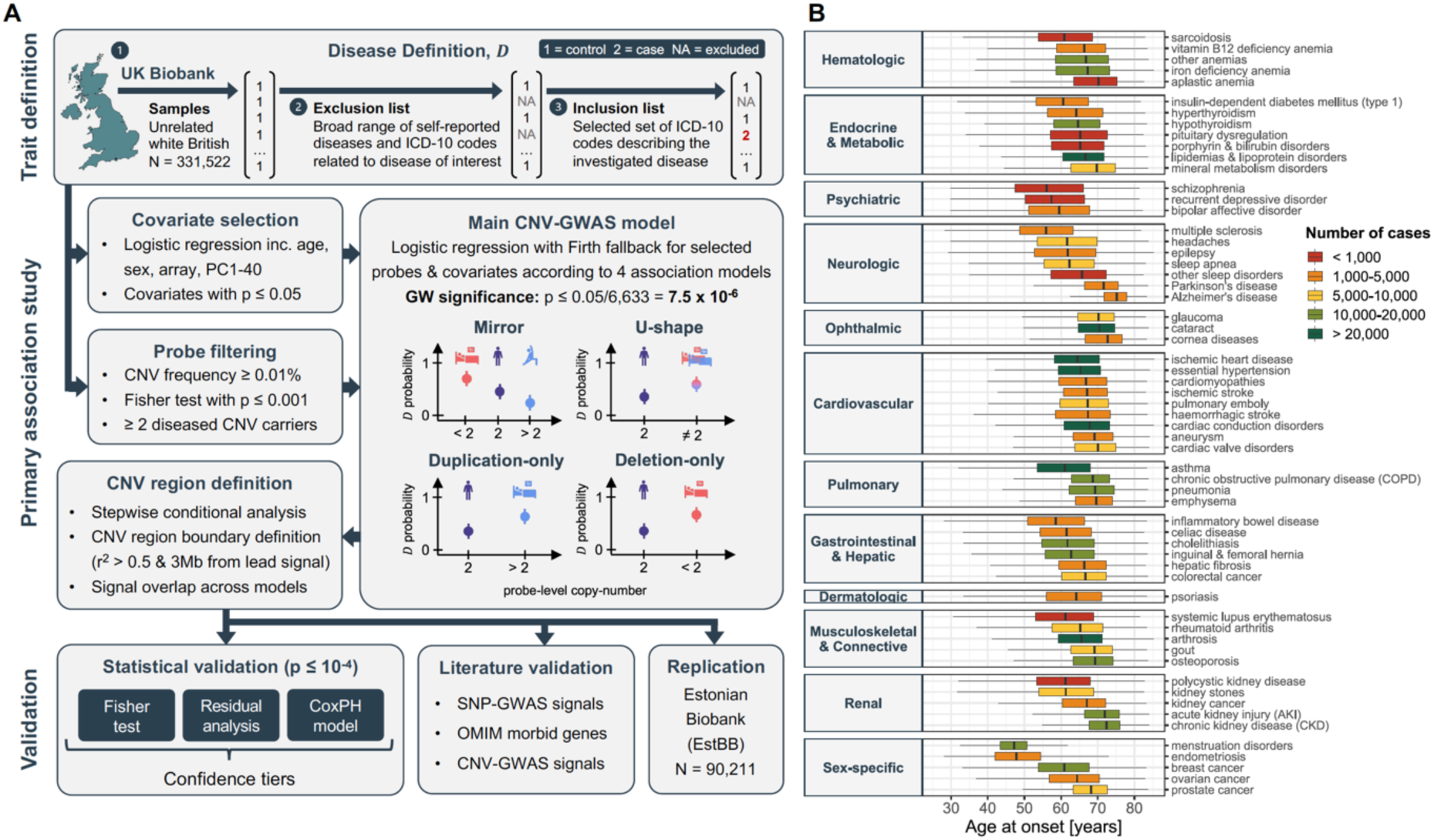
Overview of the study. (**A**) Schematic representation of the analysis workflow. Trait definition: For each of the 60 investigated diseases, unrelated white British UK Biobank participants were assigned as controls, individuals self-reporting or diagnosed with the disease of interest or a broader set of related conditions were excluded and set as missing, and individuals with a hospital-based ICD-10 diagnosis of the condition of interest were re-introduced as cases. Primary association study: Disease-specific relevant covariates were selected. Probes were pre-filtered based on copy-number variant (CNV) frequency, required to associate with the disease, and a minimum of two diseased carriers was required for the probe to be carried forward. Disease- and model-specific covariates and probes were used to generate tailored CNV genome-wide association studies (GWASs) based on Firth fallback logistic regression according to a mirror, U-shape, duplication-only (i.e., considering only duplications), and deletion-only (i.e., considering only deletions) models. Independent lead signals were identified through stepwise conditional analysis and CNV regions were defined based on probe correlation and merged across models. Validation: Statistical validation methods (i.e., Fisher test, residuals regression, and Cox proportional hazards model (CoxPH)) were used to rank associations in confidence tiers. Literature validation approaches leverage data from independent studies to corroborate that genetic perturbation (i.e., single-nucleotide polymorphisms (SNP), rare variants from the OMIM database, and CNVs) in the region are linked to the disease. Independent replication in the Estonian Biobank. (**B**) Age at onset for the 60 assessed diseases, categorized based on ICD-10 chapters and colored according to case count. Data are represented as boxplots; outliers are not shown.

### Copy-number variant genome-wide association study

To assess whether disease susceptibility is modulated by CNVs, we performed CNV genome-wide association studies (GWASs), i.e., test if the copy-number of selected probes influence the probability to develop a disease or an individual’s disease burden (i.e., number of diagnoses among the 60 studied diseases) (see Methods; Figure 1A; middle). Briefly, microarray-called CNVs for 331,522 unrelated white British individuals were transformed to the probe level after quality-control [23]. As CNVs can act through different gene dosage mechanisms, four association models were assessed: mirror and U-shape models consider deletions and duplications simultaneously, assuming that they impact disease risk in opposite or identical direction, respectively, while the CNV type-specific duplication- and deletion-only models assess independently the effect of duplications and deletions, respectively. To reduce the number and complexity of implemented logistic regressions, pre-processing steps selected relevant covariates and probes for each disease and model combination, thereby lowering computation time and decreasing the multiple testing burden (Figure S2; Table S2). All summary statistics are available (Data Availability).

Stepwise conditional analysis narrowed GW significant associations (p ≤ 7.5 x 10^-6^; see Methods for threshold calculation) to 40, 41, 21, and 38 independent signals for the mirror, U-shape, duplication-only, and deletion-only models, respectively. These were combined into 70 risk-increasing (i.e., no disease-protecting CNV) associations and 3 disease burden associations (Figure 2; Table S3). Forty-five associations (45/73 = 62%) were supported at GW significance by multiple models, the lowest p-value (i.e., “best model”) being obtained through the mirror, deletion-only, U-shape, and duplication-only models for 24, 23, 21, and 5 of the signals, respectively. No association was detected at GW significance by both the duplication-only and deletion-only models, so that each signal was attributed a “main model” that indicates whether the association is primarily driven by duplications or deletions (see Methods; Figure 2). The main model should be interpreted with caution as both deletions and duplications might influence disease risk but only one CNV type-specific model might reach GW significance (e.g., due to higher frequency). This is particularly relevant as 73% (33/45) of disease-associated CNV regions (CNVRs) have a higher duplication than deletion frequency (Figure 2A). Hence, 95% (20/21) of signals mainly driven by duplications were also identified by the mirror/U-shape model(s) and contribution of deletions cannot be excluded.

**Figure 2.**
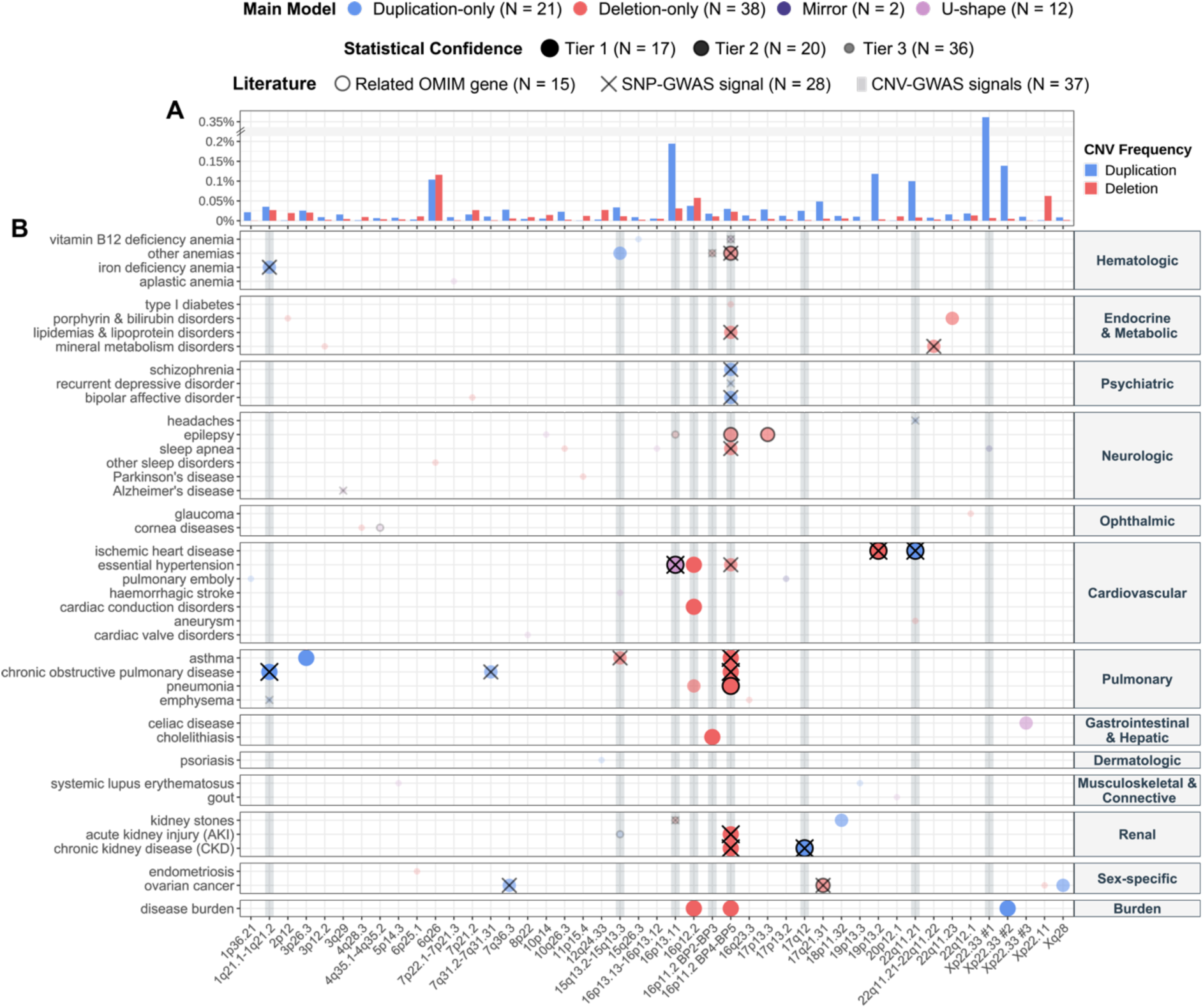
CNV-disease association map. (**A**) Duplication and deletion frequencies ([%]; y-axis; break: //) of the lead probe for each unique and non-overlapping disease-associated CNV region (CNVR), labeled with corresponding cytogenic band (x-axis; 16p11.2 is split to distinguish the distal 220kb breakpoint BP2-3 and proximal 600kb BP4-5 CNVRs; non-overlapping CNVRs on the same cytogenic band are numbered). If signals mapping to the same CNVR have different lead probes, the maximal frequency was plotted. (**B**) Associations between CNVRs (x-axis) and diseases (y-axis) identified through CNV-GWAS. Color indicates the main association model. Size and transparency reflect the statistical confidence tier. Black contours indicate overlap with OMIM gene causing a disease with shared phenotypic features. Black crosses indicate overlap with SNP-GWAS signal for a related trait. Grey shaded lines indicate CNVRs with continuous trait associations [23]. N provides count for various features.

### Validation of identified CNV-GWAS signals

Across the 45 CNVRs, CNV frequencies were low, ranging between 0.01% (our frequency cutoff) and 0.36%, with 87% (39/45) of CNVRs having a frequency ≤ 0.1% (Figure 2A). Consequently, associations rely on a low number of diseased CNV carriers and require validation (see Methods; Figure 1A; bottom; Figure 2B; Table S3). We used three statistical approaches to assess the robustness of CNV-diseases associations: i) Fisher test, ii) residual regression, and iii) time-to-event analysis through CoxPH modeling. We replicated 28/70 (40%), 23/70 (33%), and 70/70 (100%) of the associations with the respective methods at the arbitrary validation threshold of p ≤ 10^-4^. This allowed to stratify associations in confidence tiers, with 17 signals replicating with all methods (tier 1), 20 with two (tier 2), and 36 only through time-to-event analysis (tier 3). Importantly, time-to-event analysis showed that CNVs always contributed to an earlier age of disease onset, in line with the paradigm that diseases with a strong genetic etiology have earlier onset [53].

In parallel, we gathered literature evidence linking genetic variation at CNVRs with relevant phenotypes (Table S3). Forty-eight signals (48/73 = 64%) mapped to a CNVR harboring a least one OMIM morbid gene and in 15 cases, the gene was linked to a Mendelian disorder sharing phenotypic features with the associated common disease. For instance, association between 4q35 CNVs and corneal conditions (chr4:186,687,554-187,182,384; OR_U-shape_ = 18.2; 95%-CI [5.2; 63.1]; p = 5.0 x 10^-6^) encompasses *CYP4V2* [MIM: 608614], a gene associated with autosomal recessive Bietti crystalline corneoretinal dystrophy [MIM: 210370], a disorder that impairs vision and progresses to blindness by age 50-60 years [54]. We next assessed whether SNPs overlapping disease-associated CNVRs were reported to associate with the implicated disease or a biomarker thereof in the GWAS Catalog. This was the case for 28 (28/66 = 42%) autosomal signals, a similar proportion (38%) than for continuous trait CNV-GWAS [23]. For instance, distal 22q11.2 CNVs increased risk for disorders of mineral metabolism (chr22:21,797,101-22,661,627; OR_mirror_ = 0.02; 95%-CI [0.006; 0.083]; p = 9.9 x 10^-9^) and overlapped heel bone mineral density SNP-GWASs signals, while 3q29 CNVs increased Alzheimer’s disease risk (chr3:196,953,177-197,331,898; OR_U-shape_ = 11.8; 95%-CI [4.0; 34.7]; p = 6.6 x 10^-6^) and overlapped with SNP-GWAS signal for PHF-tau levels, and suggestive signals (p < 5 x 10^-6^) for frontotemporal dementia and cognitive decline in Alzheimer’s disease. Finally, 37 signals (37/73 = 51%) mapped to nine CNVRs previously found to be associated with complex traits [23].

We also set out to replicate association signals in 90,211 unrelated EstBB individuals [40], using similarly case definition than for the UKBB (see Methods; Figure S1B). Requesting at least one diseased CNV carrier, 33 of 73 associations could be evaluated, among which four were strictly replicated (p ≤ 0.05/33 = 1.5 x 10^-3^) and five additional ones reached nominal significance (p ≤ 0.05) (Table S3). Compared to what would be expected by chance, this corresponds to a 5.5-fold (p_binomial_ = 2.5 x 10^-5^) and 30.3-fold (p_binomial_ = 6.6 x 10^-7^) enrichment for replication at p ≤ 0.05 and p ≤ 5 x 10^-3^, respectively (Figure 3A). Despite low power, these results support validity of the primary UKBB association signals. Signals replicating at nominal significance are detailed in Figure 3B. Many harbor SNP-GWAS signals for related phenotypes (7/9), relevant morbid OMIM genes (4/9), or map to CNVRs previously associated with similar diseases (6/7) or biomarkers (4/9). Among them, two are in the lowest UKBB confidence tier. 15q13 duplications were linked to increased risk for acute kidney injury (AKI; chr15:30,946,160-31,881,106 | UKBB: OR_dup_ = 4.6; 95%-CI [2.5; 8.4]; p = 7.1 x 10^-7^ | EstBB: p = 2.4 x 10^-4^). Homozygous mutations in *FAN1* [MIM: 613534], one of the five genes mapping to this CNVR, have been linked to karyomegalic interstitial nephritis [MIM: 614817], a progressive renal condition that leads to CKD [55]. The second example links CNVs affecting exon 2 and intron 2-3 of *PRKN* ([MIM: 602544]) – a gene causing juvenile autosomal recessive Parkinson’s disease [MIM: 600116] – to sleep disorders such as insomnia and hypersomnia (chr6:162,705,164-162,873,489 | UKBB: OR_mirror_ = 0.12; 95%-CI [0.05; 0.26]; p = 1.6 x 10^-7^ | EstBB: p = 0.047). This finding is particularly relevant given the region’s high CNV frequency (0.22%) and the fact that sleep disturbances are among the earliest symptoms of Parkinson’s disease [56]. Follow-up studies should determine whether these individuals are more prone to develop Parkinson’s disease in the future.

**Figure 3.**
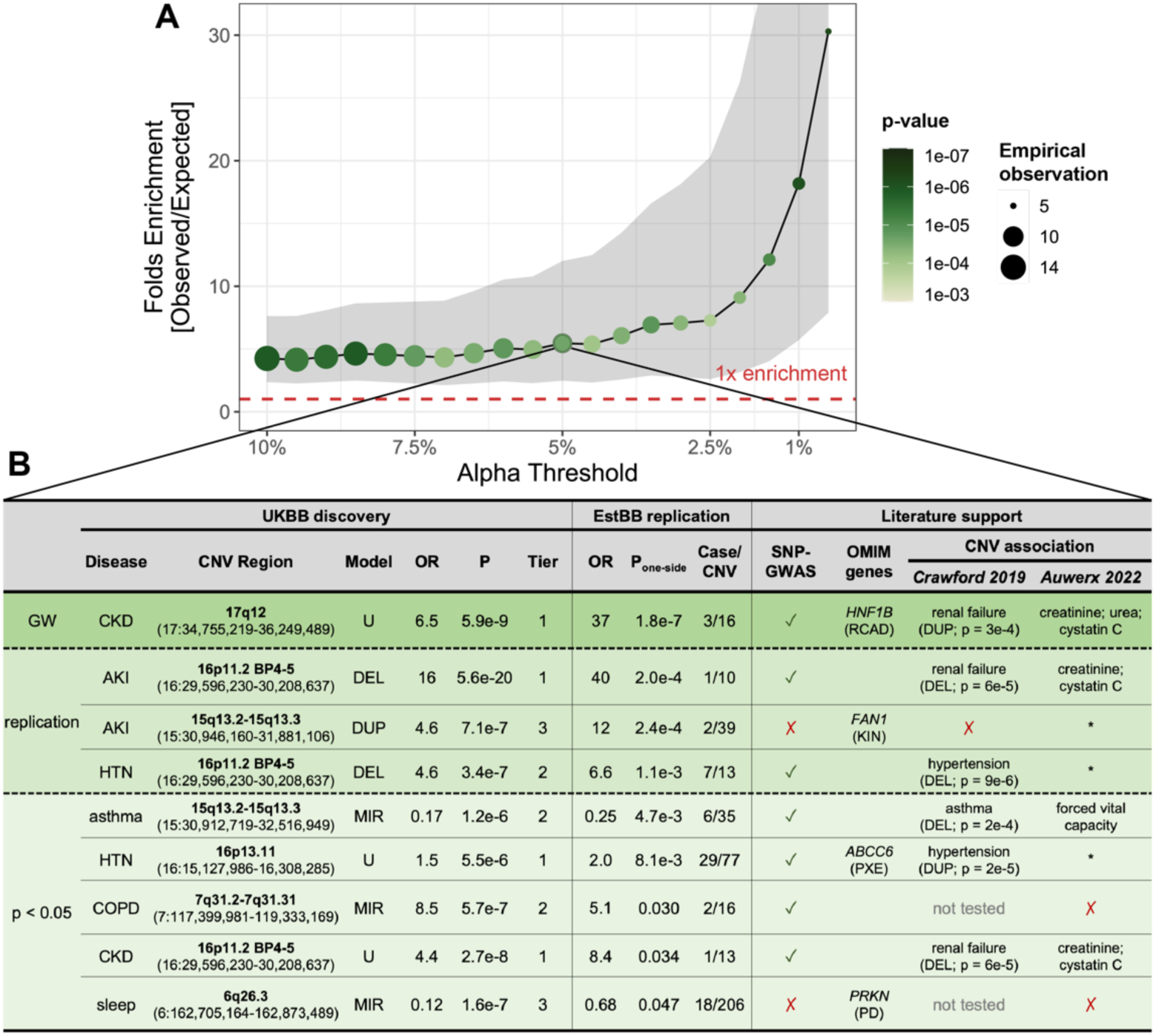
Replication of CNV-disease associations in the Estonian Biobank. (**A**) Enrichment for signal replication (y-axis; 95% confidence interval as grey ribbon) at different levels of significance (alpha; x-axis) in the Estonian Biobank (EstBB). Color and size indicate the p-value of the enrichment (one-sided binomial test) and the number of observed associations, respectively. Dashed red line indicates 1x enrichment, i.e., the number of observed associations matches the number of expected ones. (**B**) Associations replicated at nominal significance in the EstBB, color-stratified according to whether they meet the discovery genome-wide (GW; p ≤ 7.5 x 10^-6^; dark green), replication (p ≤ 1.5 x 10^-3^; green), or nominal (p ≤ 0.05; light green) significance threshold. Disease (CKD = chronic kidney disease; AKI = acute kidney injury; HTN = hypertension; COPD = chronic obstructive lung disease), cytogenic band and coordinates, best model (MIR = mirror; U = U-shape; DUP = duplication- only; DEL = deletion-only), odds ratio (OR), p-value (P) and statistical confidence tier are given for the UK Biobank (UKBB) discovery analysis. OR, one-sided p-values, and number of cases among CNV carriers are provided for the EstBB replication. Overlap with SNP-GWAS signals for a related trait (✓ = yes; ✗ = no) or a relevant OMIM gene (RCAD = renal cyst and diabetes; KIN = karyomegalic interstitial nephritis; PXE = pseudoxanthoma elasticum; PD = Parkinson’s disease) is indicated. Previous association with diseases[20] (duplication (DUP) or deletion (DEL) was associated with indicated disease; no association (✗); some CNVRs were not tested) and continuous traits [23] (disease-relevant biomarkers are specified; other traits (*); no association (✗)) are listed.

Evidence provided by statistical, literature-based, or independent replication help prioritizing the most promising associations for follow-up studies and pinpoint plausible candidate genes. We highlight several examples where deviations by one copy-number are linked to common diseases sharing clinical features with rare Mendelian conditions caused by homozygous perturbations of the same genetic region. This argues against a dichotomic view on dominant versus recessive modes of inheritance and analogously to allelic series, suggest that Mendelian and common diseases represent different ends of the phenotypic spectrum caused by genetic variation at a given locus.

### Global characterization of disease-associated CNV regions

We sought to identify global characteristics that distinguish disease-associated CNVRs (Table S4). Number of protein-coding genes embedded in disease-associated CNVRs, hereafter referred to as “disease genes”, ranged from 0 to over 30 and generally correlated with the number of encompassed probes (π_Pearson_ = 0.50; p = 4.2 x 10^-4^; Figure S3A). Exceptions include single-gene CNVRs overlapping known pathogenic genes captured thanks to high probe coverage (e.g., *BRCA1*). While only seven CNVRs (16%) associated with multiple diseases, propensity for pleiotropy depended on CNV length (+0.16 association/disease gene; p = 1.5 x 10^-5^). Accordingly, CNVRs containing more than five genes were also more likely to associate with continuous traits (OR_Fisher_ = 53.2; p = 8.5 x 10^-6^) [23]. One CNVR that stood out is the 600kb 16p11.2 BP4-5 region (Figure 2B). Originally identified as a major risk factor for autism, schizophrenia, developmental delay and intellectual disability, macro-/microcephaly, epilepsy, and obesity [57–63], we previously found the region to associate with 26 continuous complex traits [23]. Here, we show that 16p11.2 BP4-5 deletions increase the risk of 12 diseases – including both new and previously reported associations across multiple organ systems – as well as the disease burden (+3 diseases/deletion; p = 1.2 x 10^-26^), while the region’s duplication drove increased risk for psychiatric conditions (i.e., bipolar disorder, schizophrenia, and depression), in line with previous findings [62].

Next, we assessed whether disease genes were under stronger evolutionary constraint (i.e., less tolerant to mutations) than genes affected by CNVs at the same frequency but not associated with any disease (i.e., “background genes”). Compared to background genes, the 231 disease genes had more constrained pLI (p_Wilcoxon_ = 1.3 x 10^-4^; Figure S3B) and LOEUF (p_Wilcoxon_ = 1.9 x 10^-7^; Figure S3C) scores, suggesting stronger intolerance to LoF mutations. Splitting CNVRs depending on whether they have at least one association through either the duplication-only or deletion-only model, we evaluated whether embedded disease genes were sensitive to having less (i.e, haploinsufficiency; Figure S3D) or more (i.e., triplosensitivity; Figure S3E) than two functional copies. No significant difference in pHaplo scores were observed but genes overlapping regions whose duplication (p_Wilcoxon_ = 9.0 x 10^-19^) and deletion (p_Wilcoxon_ = 1.0 x 10^-23^) have been linked to diseases were more likely to be triplosensitive than background genes. Similar trends were observed considering genes overlapping CNVRs involved uniquely through the duplication-only and deletion-only models and not the other CNV type-specific model (Figure S3F-G). Overall, our results indicate that a CNVR’s pathogenicity is determined both by the number and characteristics of affected genes.

### New insights in known disease genes

Two out of 12 female *BRCA1* deletion carriers were diagnosed with ovarian cancer (chr17:41,197,733-41,276,111; OR_del_ = 284.3; 95%-CI [24.6; 3290.8]; p = 6.1 x 10^-6^; Figure 4A). *BRCA1* [MIM: 113705] is a tumor suppressor gene whose LoF represents a major genetic risk factor for the development of hereditary breast and ovarian cancer (HBOC) [MIM: 604370] [64]. Exploring the clinical records of the 12 deletion carriers, we found five diagnoses of breast cancer (a trait assessed by CNV-GWAS but that did not yield a GW-significant association), one of endometrial cancer, and one of Fallopian tube cancer, so that eight carriers (67%) had received a HBOC diagnosis (Figure 4B). Not only was prevalence of HBOC higher among *BRCA1* deletion carriers (OR_Fisher_ = 31.0; p = 1.1 x 10^-6^), but disease onset was earlier (HR = 17.0; p = 1.3 x 10^-15^; Figure 4C). Among the four carriers with no HBOC, two had received cancer prophylactic surgery, de facto reducing the penetrance of the deletion. Surgeries were likely carried out based on family history of HBOC, which was reported for 6 carriers (50%), suggesting that these deletions are inherited. We did not observe higher prevalence of other cancer types (Figure 4B).

**Figure 4.**
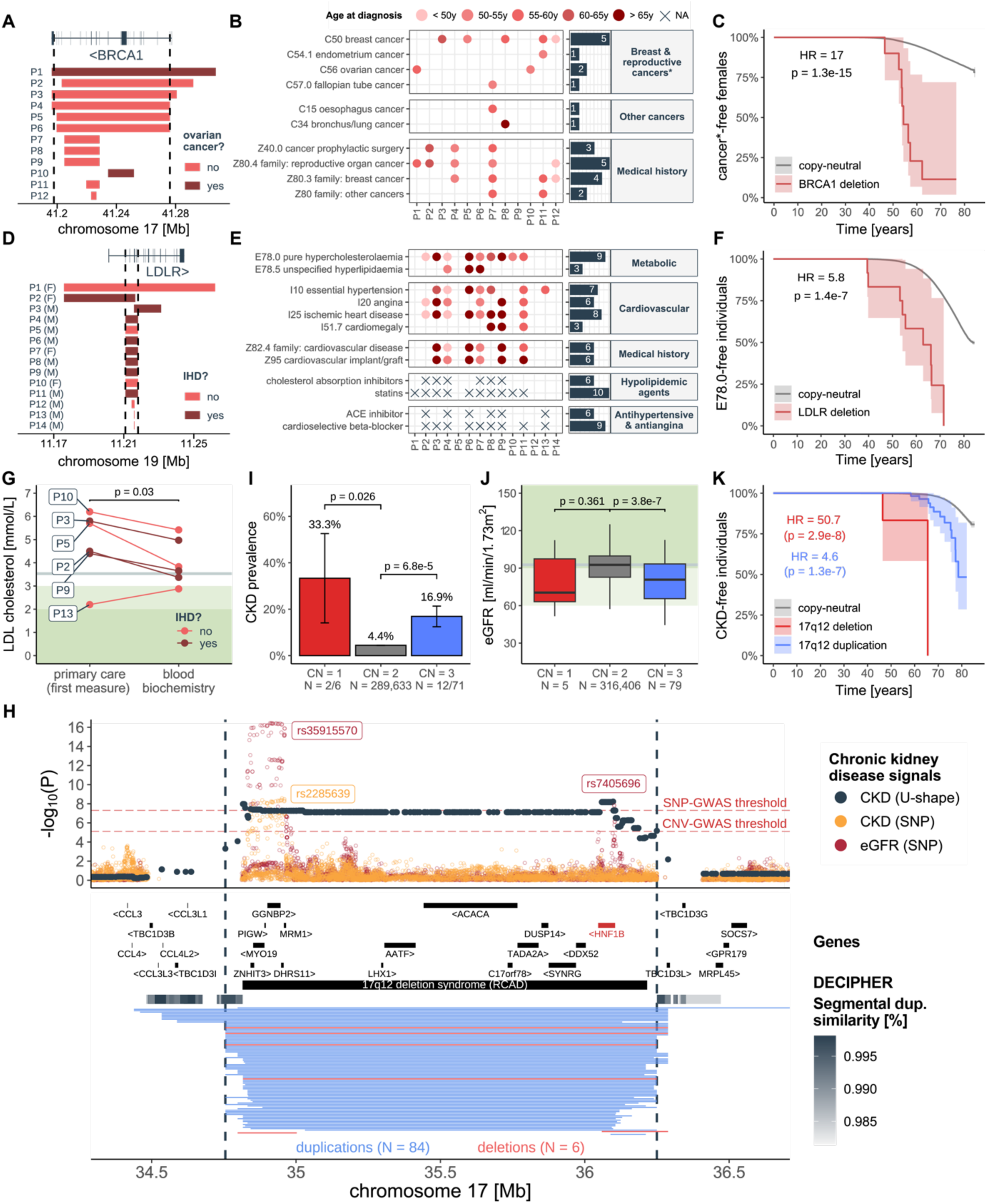
Refining contribution of CNVs to gene-disease pairs. (**A**) Genomic coordinates of the 12 females (P1-12) carrying a *BRCA1* deletion (CNVR delimited by vertical dashed lines), colored according to ovarian cancer diagnosis. (**B**) Left: Cancer and related family/personal diagnoses received by individuals in (A). Color indicates age at diagnosis. Right: Counts per ICD-10 code. (**C**) Kaplan-Meier curve depicting the percentage, with 95% confidence interval, of females free of female-specific cancers over time among copy-neutral and *BRCA1* deletion carriers. Hazard ratio (HR) and p-value for the *BRCA1* deletion are given (CoxPH model). (**D**) Genomic coordinates of the 14 individuals (P1-14) carrying an *LDLR* deletion (CNVR delimited by vertical dashed lines), colored according to ischemic heart disease (IHD) diagnosis. (**E**) Left: Medical conditions and family/personal diagnoses and medication received by ≥ 3 *LDLR* deletion carriers in (D), following legend in (B). (**F**) Kaplan-Meier curve for pure hypercholesterolemia (E78.0) among copy-neutral and *LDLR* deletion carriers, following legend as in (C). (**G**) Low density lipoprotein (LDL)-cholesterol levels (y-axis) from primary care data (first available measurement) and blood biochemistry (average over instances) for six deletion carriers in (D) with at least one antecedent primary care LDL-cholesterol measurement, colored according to IHD diagnosis. P-value compares the two data sources (paired one-sided t-test). Grey horizontal line represents median LDL-cholesterol value (from blood biochemistry) in non-carriers. Light and darker green background represent recommended target values for low (≤ 3 mmol/L) and high (≤ 1.8 mmol/L) risk individuals, respectively. (**H**) 17q12 association landscape. Top: Negative logarithm of association p-values of CNVs (dark grey; CNVR delimited by vertical dashed lines) and SNPs (orange) [70] with chronic kidney disease (CKD) and SNPs with estimated glomerular filtration rate (eGFR; red) [71]. Lead SNPs are labeled. Red horizontal dashed lines represent the genome-wide threshold for significance for CNV-GWAS (p ≤ 7.5 x 10^-6^) and SNP-GWAS (p ≤ 5 x 10^-8^). Middle: Genomic coordinates of genes and DECIPHER CNV, with *HNF1B*, the putative causal gene in red. Segmental duplications are represented as a gray gradient proportional to the degree of similarity. Bottom: Genomic coordinates of duplications (blue) and deletions (red) of UK Biobank participants overlapping the region. (**I**) CKD prevalence (± standard error) according to 17q12 copy-number (CN). P-values compare deletion (CN = 1) and duplication (CN = 3) carriers to copy-neutral (CN = 2) individuals (two-sided Fisher test). Number of cases and samples sizes are indicated (N = cases/sample size). (**J**) eGFR levels according to 17q12 CN, shown as boxplots; outliers are not shown. P-values comparisons as in (I) (two-sided t-test). Grey horizontal line represents median eGFR in non-carriers. Light and darker green background represent mildly decreased (60-90 ml/min/1.73m^2^) and normal (≥ 90 ml/min/1.73m^2^) kidney function, respectively. (**K**) Kaplan-Meier curve for CKD among 17q12 deletion and duplication carriers, following legend as in (C).

High abundance of *Alu* repeats make the low-density lipoprotein (LDL) receptor (*LDLR*) [MIM: 606945] susceptible to CNVs [65]. We found that deletion of exon 2-6 increased risk for ischemic heart disease (IHD; chr19:11,210,904-11,218,188; OR_del_ = 31.2; 95%-CI [7.1; 137.8]; p = 5.6 x 10^-6^), a condition present in 8 of 14 deletion carriers (Figure 4D). Heterozygous - and less frequently homozygous – mutations in *LDLR* represent the main genetic etiology for familial hypercholesterolemia [66], which is characterized by elevated LDL cholesterol and predisposition for adverse cardiovascular outcomes [67]. Previously identified in clinical studies of familial hypercholesterolemia [68], the CNVR implicated by our analysis specifically encompasses the ligand-binding domain of *LDLR* [66]. Confirming widespread prevalence and family history (43%) of cardiovascular diseases (Figure 4E), medical records of deletion carriers further revealed higher prevalence (OR_Fisher_ = 11.6; p = 7.9 x 10^-5^) and earlier onset (HR = 5.8; p = 1.4 x 10^-7^; Figure 4F) of pure hypercholesterolemia (E78.0), a code included in our lipidemia definition but that did not yield a signal pick-up by the CNV-GWAS. As we previously did not find the CNVR to associate with standardized blood biochemistry LDL levels [23], we hypothesized that the latter were lowered by hypolipidemic agents. Ten (71%) deletion carriers were on statins and six (43%) were additionally using cholesterol absorption inhibitors, while the remaining four did not receive a dyslipidemia or IHD diagnosis and harbored smaller deletions (i.e., P12-14; Figure 4E). We concluded that drugs likely masked genetically determined LDL levels, as shown by higher LDL levels in the first primary care measurement on record, measured prior to the standardized LDL measurement (p_t-test_ = 0.03; Figure 4G). Despite this, the recommended target of ≤ 1.8 mmol/L for high-risk individuals [69] was never met. By recovering known gene-disease pairs typically studied in clinical cohorts, we showcase how the rich phenotypic data from biobanks can generate insights into the mechanisms, epidemiology, and comorbidities of these diseases, implicating CNVs as important genetic risk factors.

### Biomarker CNV associations tag pathophysiological processes

Integration of biomarker and disease CNV-GWAS signals can identify high-confidence, clinically relevant associations. Heterozygous LoF of *HNF1B* [MIM: 189907] and 17q12 deletions cause renal cyst and diabetes (RCAD) [MIM: 137920], a severe disorder characterized by renal abnormalities and maturity-onset diabetes by the young [72,73]. While we previously showed that renal biomarkers were increased in duplication carriers [23], here, we demonstrate that both 17q12 deletions and duplications increase CKD risk (chr17:34,755,219-36,249,489; OR_U-shape_ = 6.5; 95%-CI [3.4; 12.1]; p = 5.9 x 10^-9^; Figure 4H), with a prevalence of 33.3% (p_t-test_ = 0.026) and 16.9% (p_t-test_ = 6.8 x 10^-5^) among deletion and duplication carriers respectively, versus 4.4% in copy-neutral individuals (Figure 4I). Results replicated in the EstBB (p = 1.8. x 10^-7^; Figure 3B) and are supported by 20% of CNV carriers showing signs of kidney disease based on estimated glomerular filtration rate (eGFR < 60 ml/min/1.73m^2^), compared to 2.2% in copy-neutral individuals (Figure 4J). Importantly, both 17q12 deletion and duplication lower age of CKD onset (HR ≥ 4.6; p ≥ 1.3 x 10^-7^; Figure 4K), providing strong evidence of the deleterious consequences on kidney health of altered dosage of 17q12.

Similarly, the blood pressure-increasing 16p12.2 deletion (chr16:21,946,523-22,440,319) [19,23] increased risk for hypertension (OR_del_ = 2.7; 95%-CI [1.9; 3.8]; p = 1.3 x 10^-8^) and cardiac conduction disorders (OR_del_ = 3.3; 95%-CI [2.2; 4.9]; p = 1.1 x 10^-8^), suggesting a role in cardiovascular health (Figure S4A-D). Primarily associated with developmental delay and intellectual disability [74,75] – proxied by decreased fluid intelligence (p_t-test_ = 8.7 x 10^-5^) and income (p_t-test_ = 1.4 x 10^-12^) in the UKBB (Figure S4E-F) – cardiac malformations are reported in ∼38% of clinically ascertained cases [76]. Among 193 UKBB deletion carriers, two (1%) had congenital insufficiency of the aortic valve (Q23.1), corresponding to a higher but not significantly different prevalence of cardiovascular malformations (Q20-28) than in copy-neutral individuals (OR_Fisher_ = 2.1; p = 0.251). The deletion also associated with pneumonia (OR_del_ = 3.0; 95%-CI [1.9; 4.6]; p = 5.4 x 10^-7^), coinciding with associations with decreased forced vital capacity [23] (Figure S4G-H) and peak expiratory flow [19], together demonstrating the clinical relevance of CNV-biomarker associations.

### Dissecting complex pleiotropic CNV regions

While some CNV signals converge onto the same underlying physiological processes, others tie apparently unrelated traits to the same genetic region, suggesting genuine pleiotropy. 16p13.11 harbors multiple, partially overlapping recurrent groups of CNVs allowing fine-mapping of signals to different subregions of the CNVR (Figure 5). Through different association models, the CNVR was linked to uncorrelated traits including epilepsy, kidney stones, hypertension, alkaline phosphatase (ALP), forced vital capacity, and age at menopause and menarche. We previously proposed *MARF1* as a candidate gene for the female reproductive phenotypes [23] and will focus here on the remaining traits.

**Figure 5.**
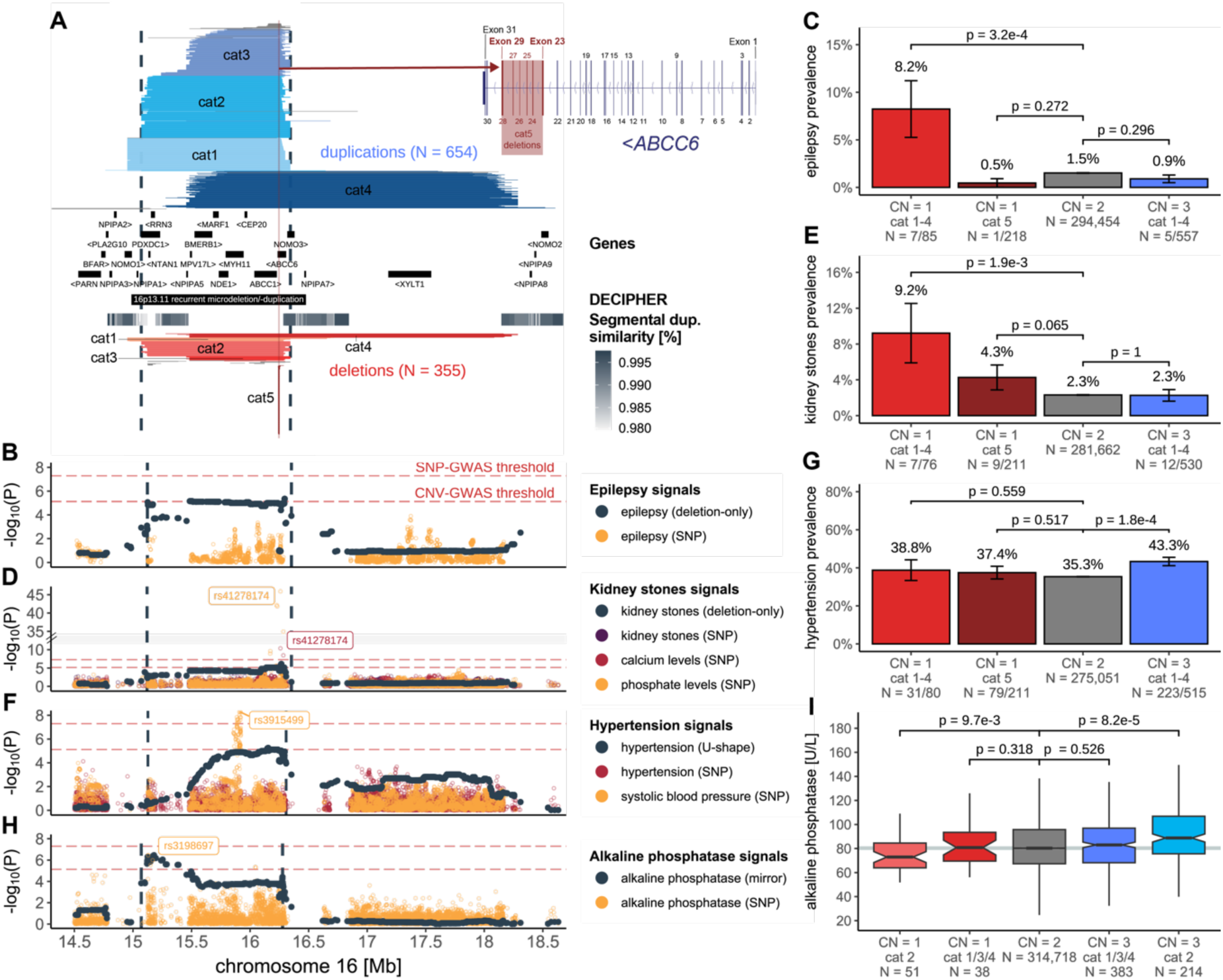
Dissection of complex pleiotropic patterns of recurrent CNVs at 16p13.11. (**A**) 16p13.11 genetic landscape. Coordinates of UK Biobank duplications (shades of blue; top) and deletions (shades of red; bottom) overlapping the maximal CNV region (CNVR delimited by vertical dashed lines) associated with epilepsy, kidney stones, hypertension, and alkaline phosphatase (ALP). CNVs are divided and colored according to five categories (cat1-5) to reflect recurrent breakpoints, with atypical CNVs in grey. Breakpoints reflect segmental duplications, represented with a grey gradient proportional to the degree of similarity. Middle: genomic coordinates of genes and DECIPHER CNV. Inset: Overlap between *ABCC6*’s exonic structure and cat5 deletions. (**B**, **D**, **F**, **H**) Negative logarithm of association p-values of CNVs with (**B**) epilepsy; (**D**) kidney stones; (**F**) hypertension; and (**H**) ALP (dark grey; model in parenthesis; CNVR delimited by vertical dashed lines) and SNPs with (**B**) epilepsy [77]; (**D**) kidney stones [78], calcium levels, and phosphate levels (y-axis; break: //); (**F**) essential hypertension and systolic blood pressure [79]; and (**H**) ALP. Lead SNPs are labeled. Red horizontal dashed lines represent genome-wide thresholds for significance for CNV-GWAS (p ≤ 7.5 x 10^-6^) and SNP-GWAS (p ≤ 5 x 10^-8^). (**C**, **E**, **G**) Prevalence (± standard error) of (**C**) epilepsy, (**E**) kidney stones, and (**G**) hypertension according to 16p13.11 copy-number (CN) and CNV categories from (A). P-values compare deletion (CN = 1) and duplication (CN = 3) carriers to copy-neutral (CN = 2) individuals (two-sided Fisher test). Number of cases and samples sizes are indicated (N = cases/sample size). (**I**) ALP levels according to 16p13.11 CN and CNV category, shown as boxplots; outliers are not shown. P-values compare deletion (CN = 1) and duplication (CN = 3) carriers to copy-neutral (CN = 2) individuals (two-sided t-test). Grey horizontal line represents median ALP value in non-carriers.

The 654 duplications and 355 deletions overlapping the maximal CNVR (chr16:15,070,916-16,353,166) were grouped into 5 categories (cat1-5) based on their breakpoints (Figure 5A; Supplemental Note 3). Risk for epilepsy was increased in deletion carriers (chr16:15,122,801-16,353,166; OR_del_ = 6.2; 95%-CI [2.8; 13.4]; p = 4.4 x 10^-6^; Figure 5B), with a prevalence of 8.3% among cat1-4 deletion carriers compared to less than 1.5% among copy-neutral and duplication carriers (Figure 5C). Previously associated with epilepsy in clinical cohorts [6,80,81], the region harbors *NDE1* [MIM: 609449], a gene associated with autosomal recessive lissencephaly [MIM: 614019] and microhydranencephaly [MIM: 605013] and whose mutation has been linked to epilepsy [82,83]. Deletions also increased risk for kidney stones (chr16:15,120,501-16,353,166; OR_del_ = 5.9; 95%-CI [2.9; 11.9]; p = 7.3 x 10^-7^), with the CNV-GWAS signal peaking close to a missense variant (rs41278174 G>A; Frequency_A_: 2.1%) in exon 23 of *ABCC6* [MIM: 603234] associating with calcium and phosphate levels through SNP-GWAS (Figure 5D). These signals coincide with the recurrent cat5 deletion that covers 29 probes spanning exons 23-29 from *ABCC6* (Figure 5A). Kidney stones prevalence reaches 4.3% among cat5 deletion carriers, in-between estimates for larger cat1-4 deletion carriers (9.3%) and copy-neutral individuals (2.3%) (Figure 5E). A wide range of variants affecting *ABCC6* have been identified and linked to the calcification disorder pseudoxanthoma elasticum through recessive [MIM: 264800] – and more rarely dominant [MIM: 177850] – inheritance [84–87], with the *Alu*-mediated cat5 deletion representing one of the most frequent variants [88,89]. *ABCC6* is expressed in the kidney and recent estimates from clinical cohorts suggested that kidney stones are an unrecognized (i.e., not used to establish clinical diagnosis) but prevalent (11-40%) feature of pseudoxanthoma elasticum [90–92]. Our data support kidney stones as a clinical outcome of *ABCC6* disruption with partial gene deletions leading to lower penetrance than large 16p13.11 deletions. Unlike epilepsy and kidney stones, both deletion (39.2%) and duplication (42%) carriers are at increased risk for hypertension (chr16:15,127,986-16,308,285; OR_U-shape_ = 1.5; 95%-CI [1.3; 1.8]; p = 5.5 x 10^-6^; Figure 5F), compared to copy-neutral individuals (35.3%) (Figure 5G). The CNVR overlaps a SNP-GWAS signal for systolic blood pressure mapping to *MYH11* [MIM: 160745] (Figure 5F). Expressed in arteries, *MYH11* encodes for smooth muscle myosin heavy chains and has been linked to dominant familial thoracic aortic aneurysm [MIM: 132900], for which hypertension represents a leading risk factor. Increased prevalence (37.4%) of hypertension among cat5 deletions implicates *ABCC6*, suggesting *cis*-epistasis for hypertension risk at 16p13.11. Consistent with this model, *ABCC6* plays a role in vascular calcification as the causal gene for generalized arterial calcification of infancy [MIM: 614473] [93,94], typically diagnosed by hypertension in newborns. Interestingly, the previously described mirror association with ALP (chr16:15,070,916-16,276,964; β_mirror_ = 6.6 U/L; p = 3.5 x 10^-7^) peaks at the distal end of the CNVR [23], nearby a suggestive SNP-GWAS signal for ALP levels (Figure 5H). Splitting ALP levels by CNV category revealed that this mirroring effect is driven by individuals with cat2 deletion (mean = 76.4 U/L; p_t-test_ = 9.7 x 10^-3^) and duplication (mean = 92.9 U/L; p_t-test_ = 8.2 x 10^-5^), as other CNV carriers had ALP levels indistinguishable from those of copy-neutral individuals (mean = 83.6 U/L) (Figure 5I). Hence, we propose the distal region of the CNVR to harbor the critical region regulating ALP levels, even if no obvious candidate gene could be identified through literature review.

The proximal 22q11.2 region, previously linked to DiGeorge [MIM: 188400] and velocardiofacial [MIM: 192430] syndromes, harbors four low-copy repeat (LCR; labeled A-to-D) [95]. Building on evidence of complex association patterns with this CNVR [38], we report novel associations between CNVs spanning LCR A-D and IHD (chr22:19,024,651-21,463,545; OR_U-shape_ = 2.1; 95%-CI [1.6; 2.8]; p = 1.5 x 10^-7^), LCR B-D and aneurysm (chr22:20,708,685-21,460,008; OR_del_ = 41.8; 95%-CI [10.0; 175.1]; p = 3.2 x 10^-7^), and LCR A-C and headaches (chr22:19,024,651-21,110,240; OR_mirror._ = 3.7; 95%-CI [2.1; 6.5]; p = 4.8 x 10^-6^) (Figure S5A; Supplemental Note 3). Based on 3 LCR B-D deletion carriers with aneurysm, this corresponds to a 22-times higher prevalence than in copy-neutral individuals (Figure S5B). Association with IHD is better powered, with a prevalence of 12%, 21%, 16%, and 20% among copy-neutral individuals and carriers of LCR C-D, B-D, and A-D CNVs, respectively (Figure S5C). This suggests that IHD risk scales with the amount of affected genetic content, supporting the presence of genetic driver(s) and/or modifier(s) in the C-D interval, beyond the prime candidate *TBX1* [MIM: 602054] [95]. Collectively, our data indicates that altered 22q11.2 dosage can result in a spectrum of cardiovascular afflictions of various degrees of severity, ranging from well-described congenital malformation [95,96] to adult-onset cardiovascular disorders.

15q13 deletions spanning BP4-5 [MIM: 612001] – and to a lesser extent duplications – have been associated with neuropsychiatric and developmental conditions [97,98], with the nicotinic acetylcholine receptor ion channel *CHRNA7* being proposed as the driver gene based on the presence of similar phenotypes in individuals with a smaller deletion (D-CHRNA7-BP5) only affecting *CHRNA7* [99] (Figure S6A). While BP4-5 duplication carriers showed higher prevalence of AKI (EstBB-replicated: Figures 3B, S6B), hemorrhagic stroke (chr15:30,912,719-31,982,408; OR_U-shape_ = 7.5; 95%-CI [3.2; 17.9]; p = 4.3 x 10^-^6; Figure S6C), and anemia (chr15:30,912,719-31,094,479; OR_dup_ = 4.9; 95%-CI [2.5; 9.7]; p = 3.2 x 10^-^6; Figure S6D), reminiscent of associations with pulse rate, mean corpuscular hemoglobin, and red blood cell count [19,23], this was not the case for the ∼10-times more numerous D-CHRNA7-BP5 duplication carriers. Replicating an association with asthma [20] (chr15:30,912,719-32,516,949; OR_mirror_ = 0.17; 95%-CI [0.08; 0.35]; p = 1.2 x 10^-6^) which parallels decreased forced vital capacity [23] and peak expiratory flow [19], this was the only deletion-driven signal for which the CNVR encompassed the entire BP4-5 region. However, only BP4-5 (46.2%; p_t-test_ = 1.8 x 10^-5^) deletion carriers had higher asthma prevalence than copy-neutral individuals (12.1%) (Figure S6E). Overall, the non-neurological disorders we associate with 15q13 CNVs appear to specifically involve dosage of the genes within BP4-D-CHRNA7 and not *CHRNA7*.

### Pathological consequences of an increased CNV burden

By assessing the global pathogenic impact of CNVs we can capture the effect of ultra-rare variants (frequency ≤ 0.01%), as well as those whose effect is not strong enough to reach GW significance under current settings. Individual-level autosomal CNV (duplication + deletion), duplication, and deletion burdens were calculated as the number of Mb or genes affected by the considered type of variation and their predictive value on the same 60 diseases (and the disease burden) previously assessed through CNV-GWAS was estimated (Figure 6A; left). Disease burden strongly associated with a high CNV load (Mb: β_CNV_ = +0.08 disease/Mb; p = 3.7 x 10^-27^ | Gene: β_del_ = +0.03 disease/gene; p = 3.7 x 10^-27^) and risk for 20 individual disorders was increased by at least one type of CNV burden (p ≤ 0.05/61 = 8.2 x 10^-4^; Figure 6B; left; Table S5). Strongest effect sizes were observed for the Mb burden for psychiatric disorders, such as bipolar disorder (OR_del_ = 1.4; p = 6.9 x 10^-4^), schizophrenia (OR_del_ = 1.4; p = 4.1 x 10^-5^), or epilepsy (OR_CNV_ = 1.1; p = 8.3 x 10^-5^), in agreement with CNVs representing important risk factors for these complex and polygenic disorders. To ensure that we do not merely capture the effect of individual CNV-disease associations previously isolated by CNV-GWAS, we excluded CNVs overlapping disease-associated CNVRs from the burden calculation and re-estimated their predictive value (Figure 6A; right). Overall association strength dropped but signal was lost only for type 1 diabetes and chronic obstructive pulmonary disease (Figure 6B; right; Table S5), suggesting that CNVs not captured by the CNV-GWAS contribute to modulating disease risk. Establishing the polygenic CNV architecture of a substantial number of common diseases, this implies that increased power will likely lead to the discovery of further CNV-disease associations.

**Figure 6.**
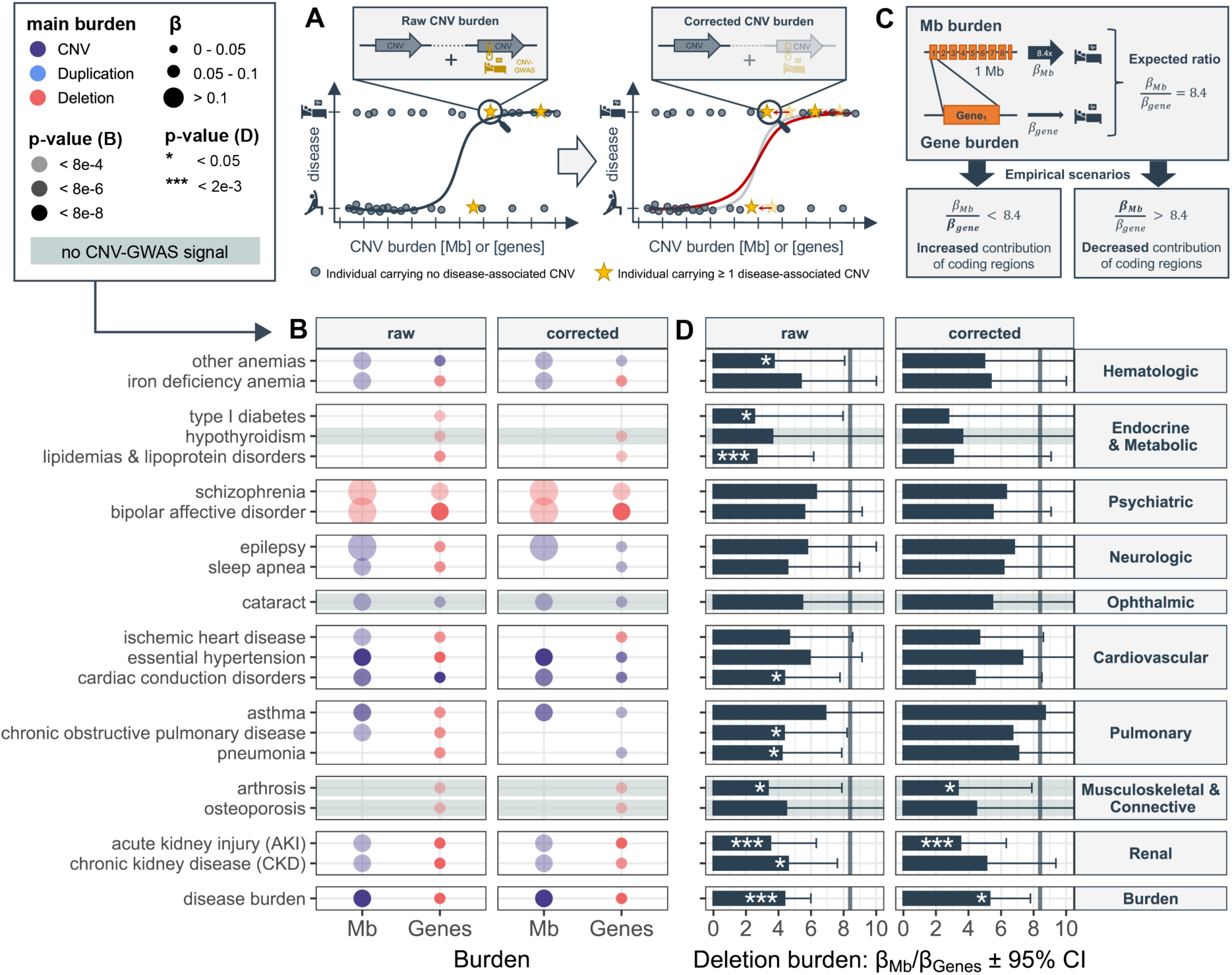
The coding deletion burden increases overall disease risk. (**A**) Burden calculation. Left: Raw CNV burden is calculated by summing up the length (in Mb or number of affected genes) of all CNVs in an individual. Burden values are used as a predictor for disease risk. Right: Corrected CNV burden is calculated by summing up the length of all CNVs that do not overlap with a CNV region (CNVR) significantly associated with the investigated disease through CNV-GWAS. Corrected burden values are used to re-estimate contribution of the CNV burden to disease risk (red curve). (**B**) Contribution of raw and corrected CNV burdens in Mb or affected genes (x-axis) to disease risk (y-axis). Only the most significantly associated burden, providing p ≤ 0.05/61 = 8.2 x 10^-4^, is shown. Color indicates whether the CNV (duplication + deletion), duplication, or deletion burden was most significantly associated, with size and transparency being proportional to the effect size (beta) and p-value, respectively. Grey horizontal bands mark traits with no CNV-GWAS signal. (**C**) Top: Based on an average genome-wide gene density of 8.4 genes/Mb, we expect the effect of the CNV burden in Mb to be 8.4x larger than the one measured in number of affected genes. Bottom: If the ratio is smaller than 8.4, it indicates an increased contribution of coding regions. Alternatively, if larger than 8.4, it indicates a decreased contribution of coding regions. (**D**) Ratio, with 95% confidence interval (CI), of the raw and corrected Mb/genes deletion burdens (x-axis) for all diseases showing at least one significant burden association. Significant deviation from the expected ratio of 8.4 (grey vertical line) is indicated as *** for p ≤ 0.05/21 = 2.4e-3 and * for p ≤ 0.05 (two-sided one sample t-test).

As the “gene burden” captures more associations than the “Mb burden” and as the deletion burden yields stronger associations than the duplication one (Figure 6B), we hypothesized that the pathogenic effect of CNVs is mainly driven by the number of deleted genes. Based on an average GW gene density of 8.4 genes/Mb, we assume the deletion burden measured in Mb (*β_Mb_*) to be 8.4 times larger than the one measured in number of affected genes (*β_gene_*), expecting a ratio of 8.4 if the type of affected genetic content does not impact disease risk (see Methods; Figure 6C). Ratios significantly (p ≤ 0.05/21 = 2.4 x 10^-3^) smaller or larger than 8.4 indicate disproportionately large or small contribution of protein coding regions, respectively. We detected a larger than expected contribution of deleted genes to the disease burden (ratio = 4.4; p = 1.3 x 10^-6^), and AKI (ratio = 3.5; p = 7.0. x 10^-4^) and lipidemia (ratio = 2.7; p = 1.4 x 10^-3^) risk, along with seven additional nominally decreased ratios (Figure 6D; left; Table S6), confirming that haploinsufficient genes mediate the burden’s effect on disease risk. Repeating the analysis with the duplication burden did not reveal any significant deviations (Table S6). After correcting for CNV-GWAS signals, only AKI (p = 7.0 x 10^-4^), arthrosis (p = 0.030), and disease burden (p = 0.016) showed increased contribution of deleted genes on disease risk, indicating that a substantial fraction of effects capture by the CNV-GWAS are mediated by deletion of coding regions.

## DISCUSSION

Using an adapted GWAS framework, we provide a detailed investigation of the contribution of CNVs to the genetic architecture of 60 common diseases and showcase how the rich phenotypic data of the UKBB can be leveraged to gain new biological insights, highlighting the role of CNVs as modulators of common disease susceptibility in the general population.

Various strategies have been used to study CNV-disease associations in the UKBB. Focusing on diseases related to the ones assessed in the current study, we replicate 10 out of the 24 detected associations (at FDR ≤ 0.1) with 54 likely pathogenic CNVs [20] and all four associations (at p ≤ 1 x 10^-9^) in a recent CNV-GWAS investigating 757 diseases [25]. Despite analyzing the same dataset, we often obtained p-values orders of magnitude smaller (e.g., 16p11.2 BP4-5 deletion and AKI: p = 5.6 x 10^-20^; p_Crawford_ = 6.0 x 10^-5^; p_Hujoel_ = 3.3 x 10^-15^). Besides accruing case count from updated hospital records, careful case-control definition and statistical handling of the binary outcomes, probe-level association analysis, and usage of different association models to mimic various dosage mechanisms could explain the increased power of our study. We consequently identified previously unreported CNV-disease associations whose relevance was asserted by follow-up analyses. Only one signal – 17q12 CNVs increasing CKD risk – was backed by all approaches, while others were supported by some analysis but not by others – e.g., two associations in the lowest statistical confidence tier were replicated in the EstBB and harbored plausible candidate genes – emphasizing the importance of considering diverse lines of corroborative evidence. Substantial overlap with relevant SNP-GWAS signals and OMIM genes indicates shared genetic mechanisms with convergent effects on the phenotype. Another line of evidence is coincidence with a disease-relevant biomarker CNV signals, e.g., four (1q21.1-2, 15q13, 16p12.2, 16p11.2 BP4-5) out of six CNVRs decreasing forced vital capacity [23] were found to increase risk for pulmonary diseases. This demonstrates that biomarkers are efficient proxies underlying (CNV-driven) pathological processes, often increasing the statistical power to detect associations due to their continuous nature. To render binary outcomes continuous, we regressed covariates out of the disease status. With the same goal, more sophisticated approaches have recently been developed that transform binary outcomes in continuous liability scores while borrowing information from age-at-disease onset, sex, and familial history [100]. Successfully applied to SNP-based GWASs, future exploration is warranted to assess their benefit in the context of CNV-GWASs. By coupling a CNV-GWAS framework designed to account for challenges linked to disease CNV association studies in population cohorts to extensive validation, we generated a list of 73 CNV-disease pairs with various levels of supporting evidence that can inform follow-up studies.

Disease-associated CNVRs harbored genes under stronger evolutionary constraint than those lacking associations and their length correlated with their propensity for pleiotropy, indicating that as previously observed [8], both the number and the nature of genes affected by CNVs influence their pathogenicity. Consequently, large multi-gene recurrent CNVs exhibited the strongest pleiotropy. A longstanding question relates to the identification of causal genes whose altered dosage drives the phenotypic alterations observed in carriers. Models with various levels of complexity have been proposed, ranging from a single driver gene to multiple driver genes modulated by epistatic interactions with other genes in the CNVR [101]. By analyzing disease prevalence in subsets of CNV carriers, association signals could be fine-mapped to narrower regions, pinpointing candidate drivers -such as *ABCC6* for kidney stones. In other cases, our data suggests that multiple subregions of the CNVR contribute to increased risk for a given disease by *cis*-epistasis, as observed for 22q11.2 and ischemic heart disease or 16p13.11 and hypertension. Interestingly, the putative driver for phenotypes originally associated with a CNVR might not be driving our newly identified associations, as shown for the 15q13 CNVR, whose non-neurological phenotypes do not appear to be linked to altered dosage of *CHRNA7*. Beyond characterizing the pleiotropic pathological consequences of recurrent CNVRs, we demonstrate that dissection of CNV-GWAS signals can fine-map associations and provide mechanistic insights into their phenotypic expression.

All CNVs increased disease risk and led to an earlier age at onset. Incorporating age at onset information has been shown to improve power to detect associations [100], and more importantly, represents the ultimate proof of clinical relevance. Indeed, many signals mapped to regions whose genetic perturbation has been reported to be pathogenic. These include associations between well-described, clinically relevant gene-disease pairs – such as *BRCA1* and *LDLR* deletions increasing the risk for ovarian cancer and IHD, respectively – but for which the role of CNVs in a large population cohort had not been previously investigated. CNVs in these genes have high penetrance but are extremely rare in the UKBB, so that association barely reached GW significance. Follow-up analyses based on the medical records, family history, medication use, and biomarkers could recapitulate additional clinical associations and establish that these deletions were most likely inherited, thereby generating insights into their role in the general population. We further show that many CNVRs previously linked to pediatric genomic disorders also increased risk for a broad spectrum of adult-onset common diseases. These associations were probably overlooked as the medical consequences in adulthood of these etiologies is often poorly characterized owing to ascertainment bias and difficulty to gather large cohorts. While awaiting validation in clinical cohorts of CNV carriers, we hope that these findings will improve clinical characterization of genomic disorders, thereby facilitating diagnosis and allowing physicians to anticipate later-onset comorbidities. For instance, we found carriers of 16p13.11 deletions affecting *ABCC6,* the causal gene for pseudoxanthoma elasticum, to be at increased risk for kidney stones, paralleling reports from clinical cohorts showing that kidney stones represent an unrecognized feature of the disease [90–92]. Awareness of this disease feature can mitigate kidney stone risk through adapted diet and sufficient water intake. Together, our results advocate for a complex model of variable CNV expressivity and penetrance that can result in a broad range of phenotypes along the rare-to-common disease spectrum and represent fertile ground for in-depth, phenome-wide studies aiming at better characterizing specific CNV regions [38].

Corroborating the deleterious impact of CNVs on an individual’s health parameters, socio-economic status, and lifespan [18,22,23,25,31,102–105], we here speculate that it acts by increasing risk for a broad range of common diseases beyond their known role in neuropsychiatric disorders [4–7]. While both duplications and deletions contributed to increased disease risk, the deletion burden’s impact was much stronger – especially for metabolic, psychiatric, and musculoskeletal diseases – and for about half the diseases, predominantly driven by deletion of coding region, in line with the commonly accepted view that deletions tend to be more deleterious than duplications and that genetic variation affecting coding regions have stronger phenotypic consequences. Only a marginal fraction of the CNV burden’s contribution to disease risk was captured by disease-associated CNVRs and risk for four diseases, i.e., hypothyroidism, cataract, arthrosis, and osteoporosis, associated with the burden despite lacking CNV-GWAS signals. With over 10,000 cases, these diseases were not underpowered compared to others, suggesting genuine differences in the genetic architecture, and illustrating the added value of burden tests. Collectively, these results predict that better powered CNV-GWAS are likely to isolate further associations currently captured by the CNV burden.

A major limitation of our study is the reliance on microarray CNV calls, which allows to assess only a fraction of the CNV landscape, i.e., mostly large CNVs or in regions with high probe coverage. We speculate that small and/or multiallelic CNVs that can only be uncovered by sequencing, will have a genetic architecture closer to the one of SNPs and indels, with higher frequencies and more subtle effect sizes, resembling those of SNPs and indels. These effects, however, are more likely tagged by common variants, limiting novel discoveries. Furthermore, by detecting more events, sequencing-based studies require adapted and more stringent significance thresholds. Still, having improved breakpoint resolution, such CNV calls are also likely to enhance fine-mapping strategies. Microarray CNV calls also exhibit high false positive rates [47]. By using stringent CNV selection criteria, we decrease the latter at the cost of decreasing power to detect true associations. This aspect is particularly relevant given that the type of CNVs we assess are rare and that the UKBB is depleted for disease cases [34], resulting in low powered GWASs. While we adopt strategies to counter the lack of power, our results are likely subject to Winner’s curse, only capturing a fraction of the strongest, possibly overestimated effects. This phenomenon might be compensated by UKBB CNV carriers being at the milder end of the clinical spectrum, leading to effect underestimation. An interesting question will be to compare effect sizes from population-based studies to those emerging from clinical cohort. In the future, longitudinal follow-up of UKBB participants will increase the number of cases – especially for late-onset diseases such as Alzheimer’s or Parkinson’ diseases – allowing better powered CNV-GWASs. Alternatively, meta-analyses can boost case number through inclusion of clinical cohorts, at the cost of poorer disease definition due to imperfect data harmonization [8]. Larger and more diverse biobanks linking genotype to phenotype data [106–108] should both validate reported associations and identify new ones.

## CONCLUSIONS

Our study provides in-depth analysis of the role of CNVs in modulating susceptibility to 60 common diseases in the general population, broadening our view on how this mutational class impacts human health. Besides describing clinically relevant and actionable associations, we illustrate how complex pleiotropic patterns can be dissected to gain new insights into the pathological mechanisms of large recurrent CNVs, providing a framework that can be applied to an even larger spectrum of diseases.

## Data Availability

All data used in this study are publicly available, as described in the methods. CNV-GWAS summary statistics (UKBB) will be deposited on the GWAS Catalog upon publication and are available upon request until then.

## SUPPLEMENTAL DATA

Supplemental data include 6 figures, 6 tables, and 3 notes.

## DECLARATIONS

### Author’s contributions

CA, AR and ZK conceived the study; CA carried out the analyses with contributions from MCS, NT, and CJC; The Estonian Biobank Research Team coordinated genotyping and sequencing data acquisition in the EstBB; MJ performed the replication study in the EstBB under the supervision of RM; ZK supervised statistical analyses; SO provided guidance for time-to-event analysis; CA drafted the manuscript and generated the figures; AR and ZK made critical revisions; All authors read, approved, and provided feedback on the final manuscript.

## Acknowledgments

We thank all biobank participants for sharing their data. UKBB and EstBB computations were performed on the JURA server (University of Lausanne) and the High-Performance Computing Center (University of Tartu), respectively.

## REFERENCES

1. Sudmant PH, Rausch T, Gardner EJ, Handsaker RE, Abyzov A, Huddleston J, et al. An integrated map of structural variation in 2,504 human genomes. Nature. 2015;526:75–81.

2. Conrad DF, Pinto D, Redon R, Feuk L, Gokcumen O, Zhang Y, et al. Origins and functional impact of copy number variation in the human genome. Nature. 2010;464:704–12.

3. Zhang F, Gu W, Hurles ME, Lupski JR. Copy Number Variationin Human Health, Disease, and Evolution. Annu Rev Genomics Hum Genet. 2009;10:451–81.

4. Sebat J, Lakshmi B, Malhotra D, Troge J, Lese-martin C, Walsh T, et al. Strong Association of De Novo Copy Number Mutations with Autism. Science (1979). 2007;316:445–9.

5. Walsh T, McClellan JM, McCarthy SE, Addington AM, Pierce SB, Cooper GM, et al. Rare structural variants disrupt multiple genes in neurodevelopmental pathways in schizophrenia. Science (1979). 2008;320:539–43.

6. Mefford HC, Muhle H, Ostertag P, von Spiczak S, Buysse K, Baker C, et al. Genome-Wide Copy Number Variation in Epilepsy: Novel Susceptibility Loci in Idiopathic Generalized and Focal Epilepsies. PLoS Genet. 2010;6:e1000962.

7. Cooper GM, Coe BP, Girirajan S, Rosenfeld JA, Vu TH, Baker C, et al. A copy number variation morbidity map of developmental delay. Nat Genet. 2011;43:838–46.

8. Collins RL, Glessner JT, Porcu E, Lepamets M, Brandon R, Lauricella C, et al. A cross-disorder dosage sensitivity map of the human genome. Cell. 2022;185:3041–3055.e25.

9. Firth H V., Richards SM, Bevan AP, Clayton S, Corpas M, Rajan D, et al. DECIPHER: Database of Chromosomal Imbalance and Phenotype in Humans Using Ensembl Resources. Am J Hum Genet. 2009;84:524–33.

10. Carvalho CMB, Lupski JR. Mechanisms underlying structural variant formation in genomic disorders. Nat Rev Genet. 2016;17:224–38.

11. Collins RL, Brand H, Karczewski KJ, Zhao X, Alföldi J, Francioli LC, et al. A structural variation reference for medical and population genetics. Nature. 2020;581:444–51.

12. Abel HJ, Larson DE, Chiang C, Das I, Kanchi KL, Layer RM, et al. Mapping and characterization of structural variation in 17,795 deeply sequenced human genomes. Nature. 2020;583:83–9.

13. Halvorsen M, Huh R, Oskolkov N, Wen J, Netotea S, Giusti-Rodriguez P, et al. Increased burden of ultra-rare structural variants localizing to boundaries of topologically associated domains in schizophrenia. Nat Commun. 2020;11:1–13.

14. Chen L, Abel HJ, Das I, Larson DE, Ganel L, Kanchi KL, et al. Association of structural variation with cardiometabolic traits in Finns. Am J Hum Genet. 2021;108:583–96.

15. Beyter D, Ingimundardottir H, Oddsson A, Eggertsson HP, Bjornsson E, Jonsson H, et al. Long-read sequencing of 3,622 Icelanders provides insight into the role of structural variants in human diseases and other traits. Nat Genet. 2021;53:779–86.

16. Babadi M, Fu JM, Lee SK, Smirnov AN, Gauthier LD, Walker M, et al. GATK-gCNV: A Rare Copy Number Variant Discovery Algorithm and Its Application to Exome Sequencing in the UK Biobank. bioRxiv. 2022;2022.08.25.504851.

17. Fitzgerald T, Birney E. CNest: A novel copy number association discovery method uncovers 862 new associations from 200,629 whole-exome sequence datasets in the UK Biobank. Cell Genomics. 2022;2:100167.

18. Kendall KM, Rees E, Escott-Price V, Einon M, Thomas R, Hewitt J, et al. Cognitive Performance Among Carriers of Pathogenic Copy Number Variants: Analysis of 152,000 UK Biobank Subjects. Biol Psychiatry. 2017;82:103–10.

19. Owen D, Bracher-Smith M, Kendall KM, Rees E, Einon M, Escott-Price V, et al. Effects of pathogenic CNVs on physical traits in participants of the UK Biobank. BMC Genomics. 2018;19:1–9.

20. Crawford K, Bracher-Smith M, Owen D, Kendall KM, Rees E, Pardiñas AF, et al. Medical consequences of pathogenic CNVs in adults: Analysis of the UK Biobank. J Med Genet. 2019;56:131–8.

21. Kendall KM, Rees E, Bracher-Smith M, Legge S, Riglin L, Zammit S, et al. Association of Rare Copy Number Variants with Risk of Depression. JAMA Psychiatry. 2019;76:818–25.

22. Macé A, Tuke MA, Deelen P, Kristiansson K, Mattsson H, Nõukas M, et al. CNV-association meta-analysis in 191,161 European adults reveals new loci associated with anthropometric traits. Nat Commun. 2017;8:1–11.

23. Auwerx C, Lepamets M, Sadler MC, Patxot M, Stojanov M, Baud D, et al. The individual and global impact of copy-number variants on complex human traits. Am J Hum Genet. 2022;109:647–68.

24. Aguirre M, Rivas MA, Priest J. Phenome-wide Burden of Copy-Number Variation in the UK Biobank. Am J Hum Genet. 2019;105:373–83.

25. Hujoel MLA, Sherman MA, Barton AR, Mukamel RE, Sankaran VG, Terao C, et al. Influences of rare copy-number variation on human complex traits. Cell. 2022;185:4233–4248.e27.

26. Sinnott-Armstrong N, Tanigawa Y, Amar D, Mars N, Benner C, Aguirre M, et al. Genetics of 35 blood and urine biomarkers in the UK Biobank. Nat Genet. 2021;53:185–94.

27. Li YR, Glessner JT, Coe BP, Li J, Mohebnasab M, Chang X, et al. Rare copy number variants in over 100,000 European ancestry subjects reveal multiple disease associations. Nat Commun. 2020;11:1–9.

28. Kopal J, Kumar K, Saltoun K, Modenato C, Moreau CA, Martin-Brevet S, et al. Rare CNVs and phenome-wide profiling highlight brain structural divergence and phenotypical convergence. Nat Hum Behav. 2023;

29. Bycroft C, Freeman C, Petkova D, Band G, Elliott LT, Sharp K, et al. The UK Biobank resource with deep phenotyping and genomic data. Nature. 2018;562:203–9.

30. Wright CF, West B, Tuke M, Jones SE, Patel K, Laver TW, et al. Assessing the Pathogenicity, Penetrance, and Expressivity of Putative Disease-Causing Variants in a Population Setting. Am J Hum Genet. 2019;104:275– 86.

31. Kingdom R, Tuke M, Wood A, Beaumont RN, Frayling TM, Weedon MN, et al. Rare genetic variants in genes and loci linked to dominant monogenic developmental disorders cause milder related phenotypes in the general population. Am J Hum Genet. 2022;109:1308–16.

32. Chen R, Shi L, Hakenberg J, Naughton B, Sklar P, Zhang J, et al. Analysis of 589,306 genomes identifies individuals resilient to severe Mendelian childhood diseases. Nat Biotechnol. 2016;34:531–8.

33. Goodrich JK, Singer-Berk M, Son R, Sveden A, Wood J, England E, et al. Determinants of penetrance and variable expressivity in monogenic metabolic conditions across 77,184 exomes. Nat Commun. 2021;12:1–15.

34. Fry A, Littlejohns TJ, Sudlow C, Doherty N, Adamska L, Sprosen T, et al. Comparison of Sociodemographic and Health-Related Characteristics of UK Biobank Participants with Those of the General Population. Am J Epidemiol. 2017;186:1026–34.

35. Falconer D. The inheritance of liability to certain diseases, estimated from the incidence among relatives. Ann Hum Genet. 1965;29:51–76.

36. Senn StephenS. Statistical issues in drug development. Wiley; 2021.

37. Mollon J, Schultz LM, Huguet G, Knowles EEM, Mathias SR, Rodrigue A, et al. Impact of Copy Number Variants and Polygenic Risk Scores on Psychopathology in the UK Biobank. Biol Psychiatry. 2023;

38. Zamariolli M, Auwerx C, Sadler MC, Graaf A Van Der, Lepik K, Schoeler T, et al. The impact of 22q11. 2 copy-number variants on human traits in the general population. Am J Hum Genet. 2023;110:1–14.

39. Giannuzzi G, Schmidt PJ, Porcu E, Willemin G, Munson KM, Nuttle X, et al. The Human-Specific BOLA2 Duplication Modifies Iron Homeostasis and Anemia Predisposition in Chromosome 16p11.2 Autism Individuals. Am J Hum Genet. 2019;105:947–58.

40. Leitsalu L, Haller T, Esko T, Tammesoo ML, Alavere H, Snieder H, et al. Cohort profile: Estonian biobank of the Estonian genome center, university of Tartu. Int J Epidemiol. 2015;44:1137–47.

41. Wu P, Gifford A, Meng X, Li X, Campbell H, Varley T, et al. Mapping ICD-10 and ICD-10-CM Codes to Phecodes: Workflow Development and Initial Evaluation. JMIR Med Inform. 2019;7:e14325.

42. Sollis E, Mosaku A, Abid A, Buniello A, Cerezo M, Gil L, et al. The NHGRI-EBI GWAS Catalog: knowledgebase and deposition resource. Nucleic Acids Res. 2023;51:D977–85.

43. Hamosh A, Scott AF, Amberger JS, Bocchini CA, McKusick VA. Online Mendelian Inheritance in Man (OMIM), a knowledgebase of human genes and genetic disorders. Nucleic Acids Res. 2005;33.

44. Karczewski KJ., Francioli LC, Tiao G, Cummings BB, Alföldi J, Wang Q, et al. The mutational constraint spectrum quantified from variation in 141,456 humans. Nature. 2020;581:434–43.

45. Aguet F, Barbeira A, Bonazzola R, Brown A, Castel S, Jo B, et al. The GTEx Consortium atlas of genetic regulatory effects across human tissues. Science (1979). 2020;369:1318–30.

46. Wang K, Li M, Hadley D, Liu R, Glessner J, Grant SFA, et al. PennCNV: An integrated hidden Markov model designed for high-resolution copy number variation detection in whole-genome SNP genotyping data. Genome Res. 2007;17:1665–74.

47. Macé A, Tuke MA, Beckmann JS, Lin L, Jacquemont S, Weedon MN, et al. New quality measure for SNP array based CNV detection. Bioinformatics. 2016;32:3298–305.

48. Chang CC, Chow CC, Tellier LCAM, Vattikuti S, Purcell SM, Lee JJ. Second-generation PLINK: Rising to the challenge of larger and richer datasets. Gigascience. 2015;4:1–16.

49. Wang K, Li M, Hakonarson H. ANNOVAR: functional annotation of genetic variants from high-throughput sequencing data. Nucleic Acids Res. 2010;38:e164–e164.

50. Kent WJ, Sugnet CW, Furey TS, Roskin KM, Pringle TH, Zahler AM, et al. The human genome browser at UCSC. Genome Res. 2002;12:996–1006.

51. Gao X, Starmer J, Martin ER. A multiple testing correction method for genetic association studies using correlated single nucleotide polymorphisms. Genet Epidemiol. 2008;32:361–9.

52. Therneau TM. Survival Analysis [R package survival version 3.5-3]. 2022 [cited 2023 Apr 17]; Available from: https://CRAN.R-project.org/package=survival

53. Berman JJ. Rare Diseases and Orphan Drugs: Keys to Understanding and Treating the Common Diseases. Elsevier Inc.; 2014.

54. Li A, Jiao X, Munier FL, Schorderet DF, Yao W, Iwata F, et al. Bietti Crystalline Corneoretinal Dystrophy Is Caused by Mutations in the Novel Gene CYP4V2. Am J Hum Genet. 2004;74:817–26.

55. Zhou W, Otto EA, Cluckey A, Airik R, Hurd TW, Chaki M, et al. FAN1 mutations cause karyomegalic interstitial nephritis, linking chronic kidney failure to defective DNA damage repair. Nat Genet. 2012;44:910–5.

56. Kryger MH, Roth T, Goldstein CA. Principles and Practice of Sleep Medicine. Elsevier Health Sciences; 2021.

57. Shinawi M, Liu P, Kang SHL, Shen J, Belmont JW, Scott DA, et al. Recurrent reciprocal 16p11.2 rearrangements associated with global developmental delay, behavioural problems, dysmorphism, epilepsy, and abnormal head size. J Med Genet. 2010;47:332–41.

58. Weiss LA, Shen Y, Korn JM, Arking DE, Miller DT, Fossdal R, et al. Association between Microdeletion and Microduplication at 16p11.2 and Autism. N Engl J Med. 2008;358:667–75.

59. D’Angelo D, Lebon S, Chen Q, Martin-Brevet S, Snyder LAG, Hippolyte L, et al. Defining the Effect of the 16p11.2 Duplication on Cognition, Behavior, and Medical Comorbidities. JAMA Psychiatry. 2016;73:20–30.

60. Reinthaler EM, Lal D, Lebon S, Hildebrand MS, Dahl HHM, Regan BM, et al. 16p11.2 600 kb Duplications confer risk for typical and atypical Rolandic epilepsy. Hum Mol Genet. 2014;23:6069–80.

61. Jacquemont S, Reymond A, Zufferey F, Harewood L, Walters RG, Kutalik Z, et al. Mirror extreme BMI phenotypes associated with gene dosage at the chromosome 16p11.2 locus. Nature. 2011;478:97–102.

62. McCarthy SE, Makarov V, Kirov G, Addington AM, McClellan J, Yoon S, et al. Microduplications of 16p11.2 are associated with schizophrenia. Nat Genet. 2009;41:1223–7.

63. Walters RG, Jacquemont S, Valsesia A, De Smith AJ, Martinet D, Andersson J, et al. A new highly penetrant form of obesity due to deletions on chromosome 16p11.2. Nature. 2010;463:671–5.

64. Miki Y, Swensen J, Shattuck-Eidens D, Futreal PA, Harshman K, Tavtigian S, et al. A strong candidate for the breast and ovarian cancer susceptibility gene BRCA1. Science (1979). 1994;266:66–71.

65. Iacocca MA, Hegele RA. Role of DNA copy number variation in dyslipidemias. Curr Opin Lipidol. 2018;29:125– 32.

66. Hobbs HH, Russell DW, Brown MS, Goldstein JL. The LDL receptor locus in familial hypercholesterolemia: mutational analysis of a membrane protein. Annu Rev Genet. 1990;24:133–70.

67. Defesche JC, Gidding SS, Harada-Shiba M, Hegele RA, Santos RD, Wierzbicki AS. Familial hypercholesterolaemia. Nat Rev Dis Primers. 2017;3:17093.

68. Iacocca MA, Wang J, Dron JS, Robinson JF, McIntyre AD, Cao H, et al. Use of next-generation sequencing to detect LDLR gene copy number variation in familial hypercholesterolemia. J Lipid Res. 2017;58:2202–9.

69. Mach F, Baigent C, Catapano AL, Koskinas KC, Casula M, Badimon L, et al. 2019 ESC/EAS Guidelines for the management of dyslipidaemias: lipid modification to reduce cardiovascular riskThe Task Force for the management of dyslipidaemias of the European Society of Cardiology (ESC) and European Atherosclerosis Society (EAS). Eur Heart J. 2020;41:111–88.

70. Wuttke M, Li Y, Li M, Sieber KB, Feitosa MF, Gorski M, et al. A catalog of genetic loci associated with kidney function from analyses of a million individuals. Nat Genet. 2019;51:957–72.

71. Stanzick KJ, Li Y, Schlosser P, Gorski M, Wuttke M, Thomas LF, et al. Discovery and prioritization of variants and genes for kidney function in >1.2 million individuals. Nat Commun. 2021;12:1–17.

72. Fajans SS, Bell GI, Polonsky KS. Molecular mechanisms and clinical pathophysiology of maturity-onset diabetes of the young. N Engl J Med. 2001;345:97180.

73. Mefford HC, Clauin S, Sharp AJ, Moller RS, Ullmann R, Kapur R, et al. Recurrent Reciprocal Genomic Rearrangements of 17q12 Are Associated with Renal Disease, Diabetes, and Epilepsy. Am J Hum Genet. 2007;81:1057–69.

74. Girirajan S, Rosenfeld JA, Cooper GM, Antonacci F, Siswara P, Itsara A, et al. A recurrent 16p12.1 microdeletion supports a two-hit model for severe developmental delay. Nat Genet. 2010;42:203–9.

75. Stefansson H, Meyer-Lindenberg A, Steinberg S, Magnusdottir B, Morgen K, Arnarsdottir S, et al. CNVs conferring risk of autism or schizophrenia affect cognition in controls. Nature. 2013;505:361–6.

76. Girirajan S, Pizzo L, Moeschler J, Rosenfeld J. 16p12.2 Recurrent Deletion. GeneReviews® [Internet]. 2018 [cited 2023 Apr 4]; Available from: https://www.ncbi.nlm.nih.gov/books/NBK274565/

77. Stevelink R, Campbell C, Chen S, The International League Against Epilepsy Consortium on Complex Epilepsies. Genome-wide meta-analysis of over 29,000 people with epilepsy reveals 26 loci and subtype-specific genetic architecture. medRxiv. 2022;2022.06.08.22276120.

78. Howles SA, Wiberg A, Goldsworthy M, Bayliss AL, Gluck AK, Ng M, et al. Genetic variants of calcium and vitamin D metabolism in kidney stone disease. Nat Commun. 2019;10:1–10.

79. Evangelou E, Warren HR, Mosen-Ansorena D, Mifsud B, Pazoki R, Gao H, et al. Genetic analysis of over 1 million people identifies 535 new loci associated with blood pressure traits. Nat Genet. 2018;50:1412–25.

80. De Kovel CGF, Trucks H, Helbig I, Mefford HC, Baker C, Leu C, et al. Recurrent microdeletions at 15q11.2 and 16p13.11 predispose to idiopathic generalized epilepsies. Brain. 2010;133:23–32.

81. Heinzen EL, Radtke RA, Urban TJ, Cavalleri GL, Depondt C, Need AC, et al. Rare Deletions at 16p13.11 Predispose to a Diverse Spectrum of Sporadic Epilepsy Syndromes. Am J Hum Genet. 2010;86:707–18.

82. Alkuraya FS, Cai X, Emery C, Mochida GH, Al-Dosari MS, Felie JM, et al. Human Mutations in NDE1 Cause Extreme Microcephaly with Lissencephaly. Am J Hum Genet. 2011;88:536–47.

83. Bakircioglu M, Carvalho OP, Khurshid M, Cox JJ, Tuysuz B, Barak T, et al. The Essential Role of Centrosomal NDE1 in Human Cerebral Cortex Neurogenesis. Am J Hum Genet. 2011;88:523–35.

84. Ringpfeil F, Lebwohl MG, Christiano AM, Uitto J. Pseudoxanthoma elasticum: Mutations in the MRP6 gene encoding a transmembrane ATP-binding cassette (ABC) transporter. Proc Natl Acad Sci U S A. 2000;97:6001– 6.

85. Struk B, Cai L, Zäch S, Ji W, Chung J, Lumsden A, et al. Mutations of the gene encoding the transmembrane transporter protein ABC-C6 cause pseudoxanthoma elasticum. J Mol Med. 2000;78:282–6.

86. Le Saux O, Urban Z, Tschuch C, Csiszar K, Bacchelli B, Quaglino D, et al. Mutations in a gene encoding an ABC transporter cause pseudoxanthoma elasticum. Nat Genet. 2000;25:223–7.

87. Bergen AAB, Plomp AS, Schuurman EJ, Terry S, Breuning M, Dauwerse H, et al. Mutations in ABCC6 cause pseudoxanthoma elasticum. Nat Genet. 2000;25:228–31.

88. Le Saux O, Beck K, Sachsinger C, Silvestri C, Treiber C, Goöring HHH, et al. A Spectrum of ABCC6 Mutations Is Responsible for Pseudoxanthoma Elasticum. Am J Hum Genet. 2001;69:749–64.

89. Ringpfeil F, Nakano A, Uitto J, Pulkkinen L. Compound Heterozygosity for a Recurrent 16.5-kb Alu-Mediated Deletion Mutation and Single-Base-Pair Substitutions in the ABCC6 Gene Results in Pseudoxanthoma Elasticum. Am J Hum Genet. 2001;68:642–52.

90. Ralph D, Allawh R, Terry IF, Terry SF, Uitto J, Li QL. Kidney Stones Are Prevalent in Individuals with Pseudoxanthoma Elasticum, a Genetic Ectopic Mineralization Disorder. Int J Dermatol Venereol. 2020;3:198– 204.

91. Legrand A, Cornez L, Samkari W, Mazzella JM, Venisse A, Boccio V, et al. Mutation spectrum in the ABCC6 gene and genotype–phenotype correlations in a French cohort with pseudoxanthoma elasticum. Genetics in Medicine. 2017;19:909–17.

92. Letavernier E, Kauffenstein G, Huguet L, Navasiolava N, Bouderlique E, Tang E, et al. ABCC6 deficiency promotes development of randall plaque. Journal of the American Society of Nephrology. 2018;29:2337–47.

93. Nitschke Y, Baujat G, Botschen U, Wittkampf T, Du Moulin M, Stella J, et al. Generalized Arterial Calcification of Infancy and Pseudoxanthoma Elasticum Can Be Caused by Mutations in Either ENPP1 or ABCC6. Am J Hum Genet. 2012;90:25–39.

94. Le Boulanger G, Labrèze C, Croué A, Schurgers LJ, Chassaing N, Wittkampf T, et al. An unusual severe vascular case of pseudoxanthoma elasticum presenting as generalized arterial calcification of infancy. Am J Med Genet A. 2010;152A:118–23.

95. McDonald-McGinn DM, Sullivan KE, Marino B, Philip N, Swillen A, Vorstman JAS, et al. 22q11.2 deletion syndrome. Nat Rev Dis Primers. 2015;1:1–19.

96. Bartik LE, Hughes SS, Tracy M, Feldt MM, Zhang L, Arganbright J, et al. 22q11.2 duplications: Expanding the clinical presentation. Am J Med Genet A. 2022;188:779–87.

97. Sharp AJ, Mefford HC, Li K, Baker C, Skinner C, Stevenson RE, et al. A recurrent 15q13.3 microdeletion syndrome associated with mental retardation and seizures. Nat Genet. 2008;40:322–8.

98. Lowther C, Costain G, Stavropoulos DJ, Melvin R, Silversides CK, Andrade DM, et al. Delineating the 15q13.3 microdeletion phenotype: a case series and comprehensive review of the literature. Genetics in Medicine. 2015;17:149–57.

99. Gillentine MA, Schaaf CP. The Human Clinical Phenotypes of Altered CHRNA7 Copy Number. Biochem Pharmacol. 2015;97:352.

100. Pedersen EM, Agerbo E, Plana-Ripoll O, Grove J, Dreier JW, Musliner KL, et al. Accounting for age of onset and family history improves power in genome-wide association studies. Am J Hum Genet. 2022;109:417–32.

101. Golzio C, Katsanis N. Genetic architecture of reciprocal CNVs. Curr Opin Genet Dev. 2013;23:240–8.

102. Männik K, Mägi R, Macé A, Cole B, Guyatt AL, Shihab HA, et al. Copy number variations and cognitive phenotypes in unselected populations. JAMA. 2015;313:2044–54.

103. Wheeler E, Huang N, Bochukova EG, Keogh JM, Lindsay S, Garg S, et al. Genome-wide SNP and CNV analysis identifies common and low-frequency variants associated with severe early-onset obesity. Nat Genet. 2013;45:513–7.

104. Dauber A, Yu Y, Turchin MC, Chiang CW, Meng YA, Demerath EW, et al. Genome-wide association of copy-number variation reveals an association between short stature and the presence of low-frequency genomic deletions. Am J Hum Genet. 2011;89:751–9.

105. Saarentaus EC, Havulinna AS, Mars N, Ahola-Olli A, Kiiskinen TTJ, Partanen J, et al. Polygenic burden has broader impact on health, cognition, and socioeconomic outcomes than most rare and high-risk copy number variants. Mol Psychiatry. 2021;26:4884–95.

106. Denny JC, Rutter JL, Goldstein DB, Philippakis A, Smoller JW, Jenkins G, et al. The ‘All of Us’ Research Program. N Engl J Med. 2019;381:668–76.

107. Hunter-Zinck H, Shi Y, Li M, Gorman BR, Ji SG, Sun N, et al. Genotyping Array Design and Data Quality Control in the Million Veteran Program. Am J Hum Genet. 2020;106:535–48.

108. Kurki MI, Karjalainen J, Palta P, Sipilä TP, Kristiansson K, Donner KM, et al. FinnGen provides genetic insights from a well-phenotyped isolated population. Nature. 2023;613:508–18.

