## Supplemental Figures & Notes for "Copy-number variants as modulators of common disease susceptibility"

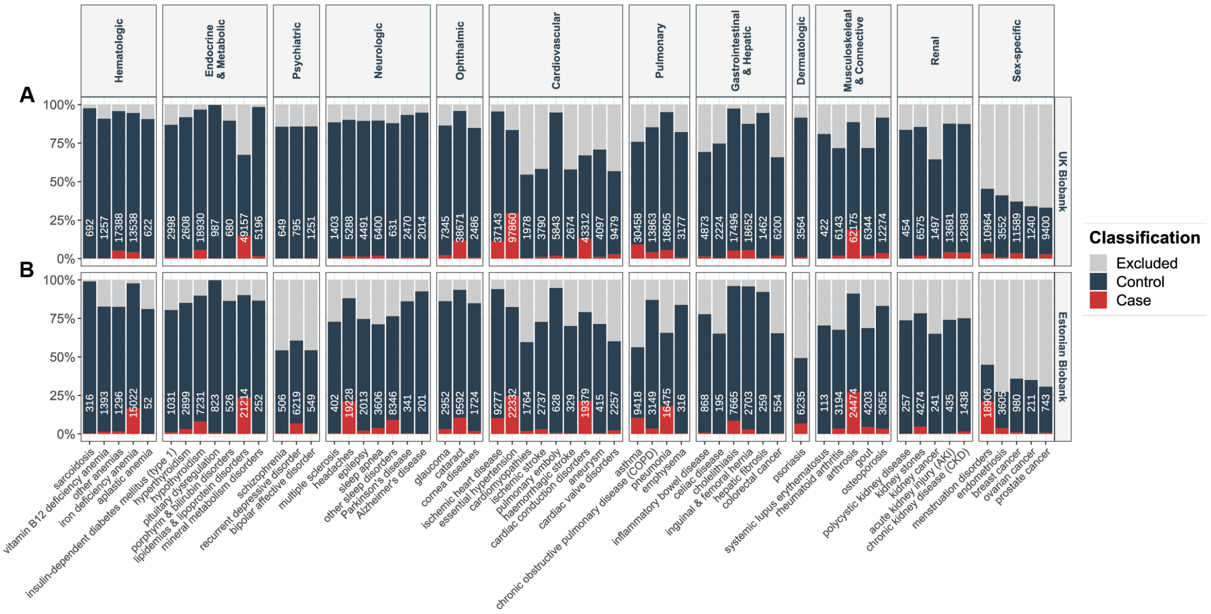


**Figure S1. Case-control distribution in the UK and Estonian Biobanks**

(**A**) UK Biobank and (**B**) Estonian Biobank percent stacked bar chart of cases (red), controls (dark blue), and excluded (grey) individuals, for each of the 60 assessed diseases categorized according to their ICD-10 chapter. Case count is indicated in white on each bar.

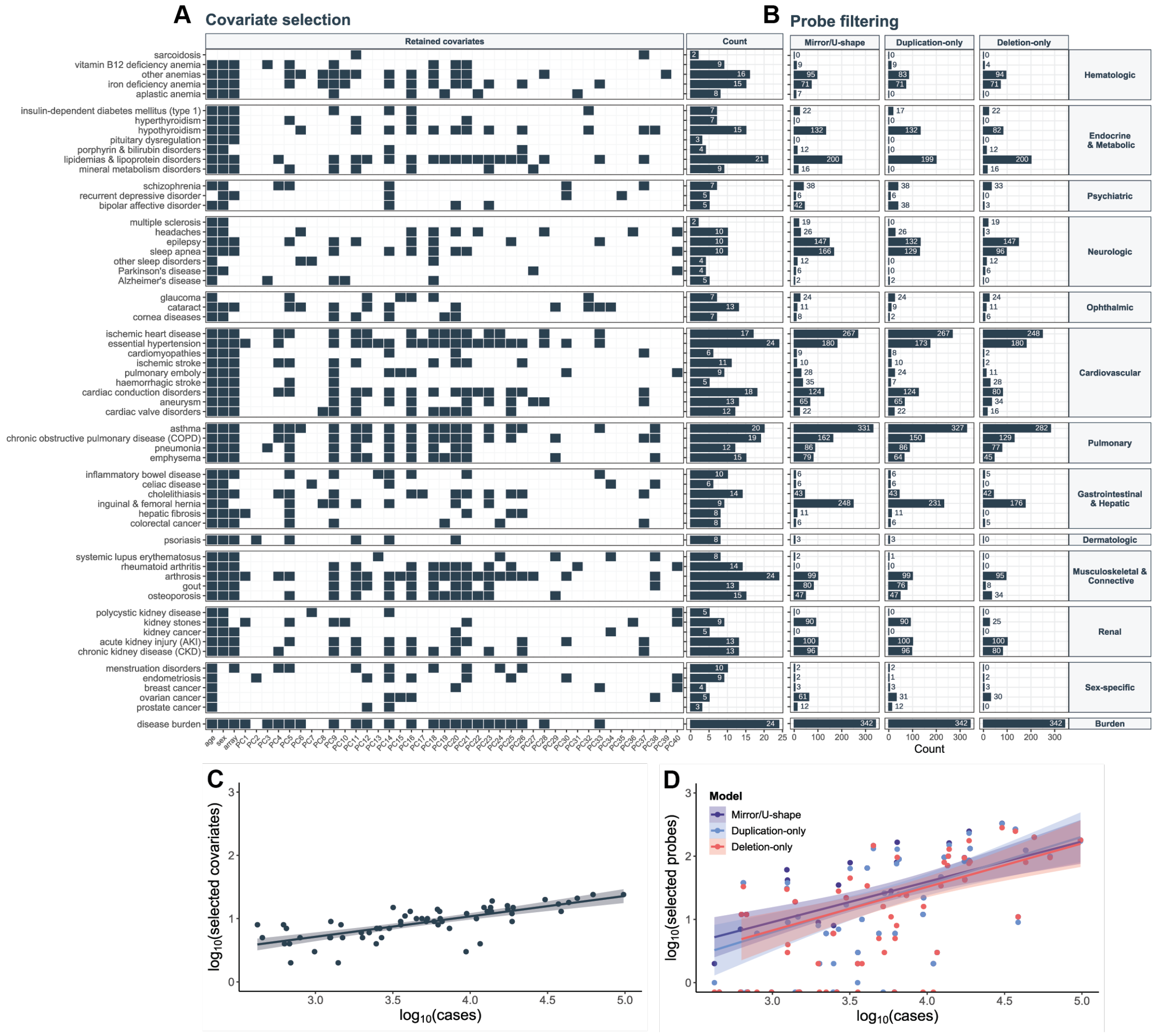


#### Figure S2. Covariate selection and probe filtering

(**A**) Left: Dark grey tiles indicate covariates (x-axis) retained for the corresponding disease and/or disease burden (y-axis) (nominal significant association). PC = principal component. Right: Number of retained covariates per disease. (**B**) Number of probes retained after filtering (x-axis) for the mirror and U-shape (left), duplication-only (middle), and deletion-only (right) models for each of the 60 investigated diseases and the disease burden (y-axis). (**C**, **D**) Logarithm of number of selected (**C**) covariates or (**D**) probes (y-axis) against the logarithm of number of cases (x-axis) for each of the 60 assessed diseases and disease burden. Linear regression equations with 95% confidence intervals are displayed. For (D), data points and equations are represented separately for the different models. Number or retained covariates (range: 2-24) and probes (range: 0-342) correlated with the case number, aligning with the increased power expected for prevalent diseases.

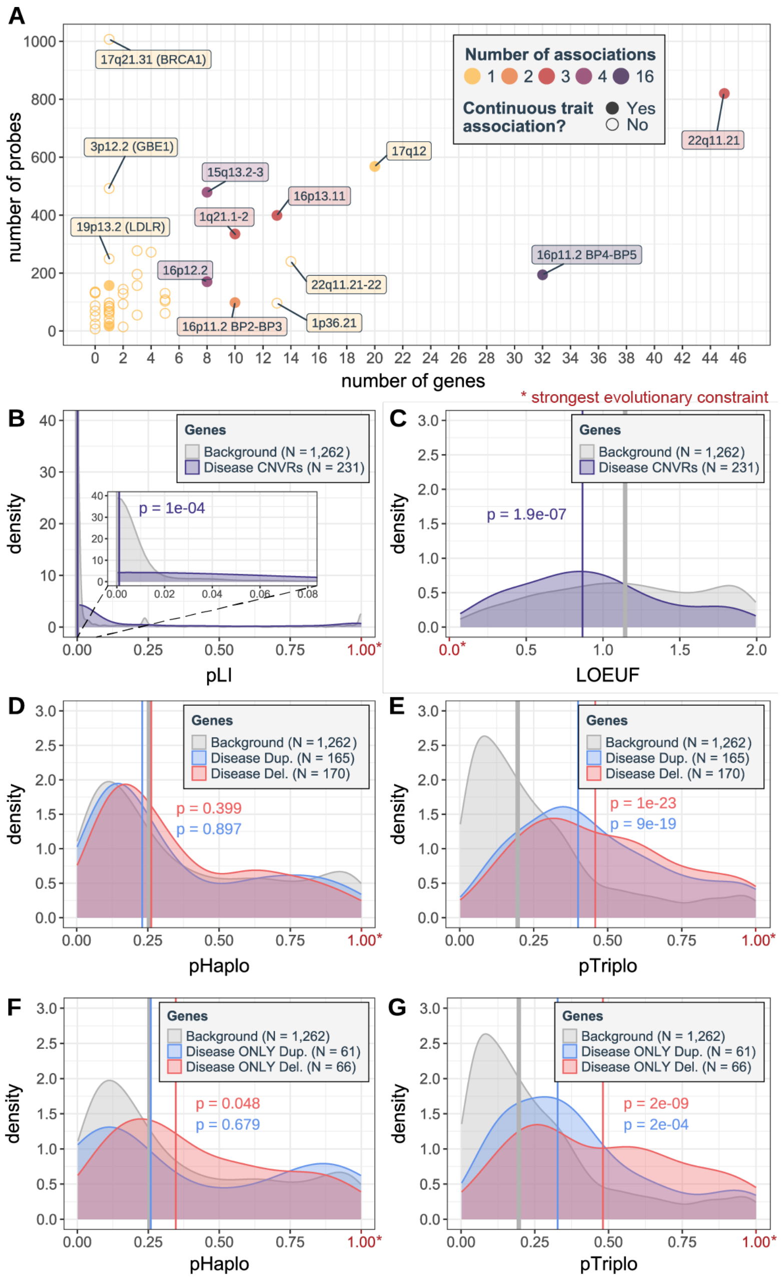


#### Figure S3. Characteristics of disease-associated CNV regions

(**A**) Number of probes (y-axis) versus number of affected genes (x-axis) for disease-associated CNV regions (CNVRs). Color reflects number of associations, with full circles indicating previous association with continuous traits [1]. CNVRs affecting ≥ 6 genes or single-gene CNVR affecting > 200 probes are labeled with cytogenic bands. (**B**-**G**) Evolutionary constraint of CNVR-encompassed genes (i.e., “disease genes”): Distribution of (**B**) pLI and (**C**) LOEUF scores for disease genes versus background genes (i.e., genes overlapping regions with a CNV frequency ≥ 0.01% but no disease association). Distribution of probability of (**D**) haploinsufficiency (pHaplo) and (**E**) triplosensitivity (pTriplo) scores for genes overlapping CNVRs significantly associated to a disease through the duplication-only or deletion-only models versus background genes. Distribution of probability of (**F**) pHaplo and (**G**) pTriplo scores for genes overlapping CNVRs *uniquely* associated to a disease through the duplication-only or deletion-only model versus background genes. Number of genes (N) and the median score (vertical line) is indicated for each group. P-values compare groups versus background gene medians (two-sided Wilcoxon test). Strongest evolutionary constraint is indicated in red with a star.

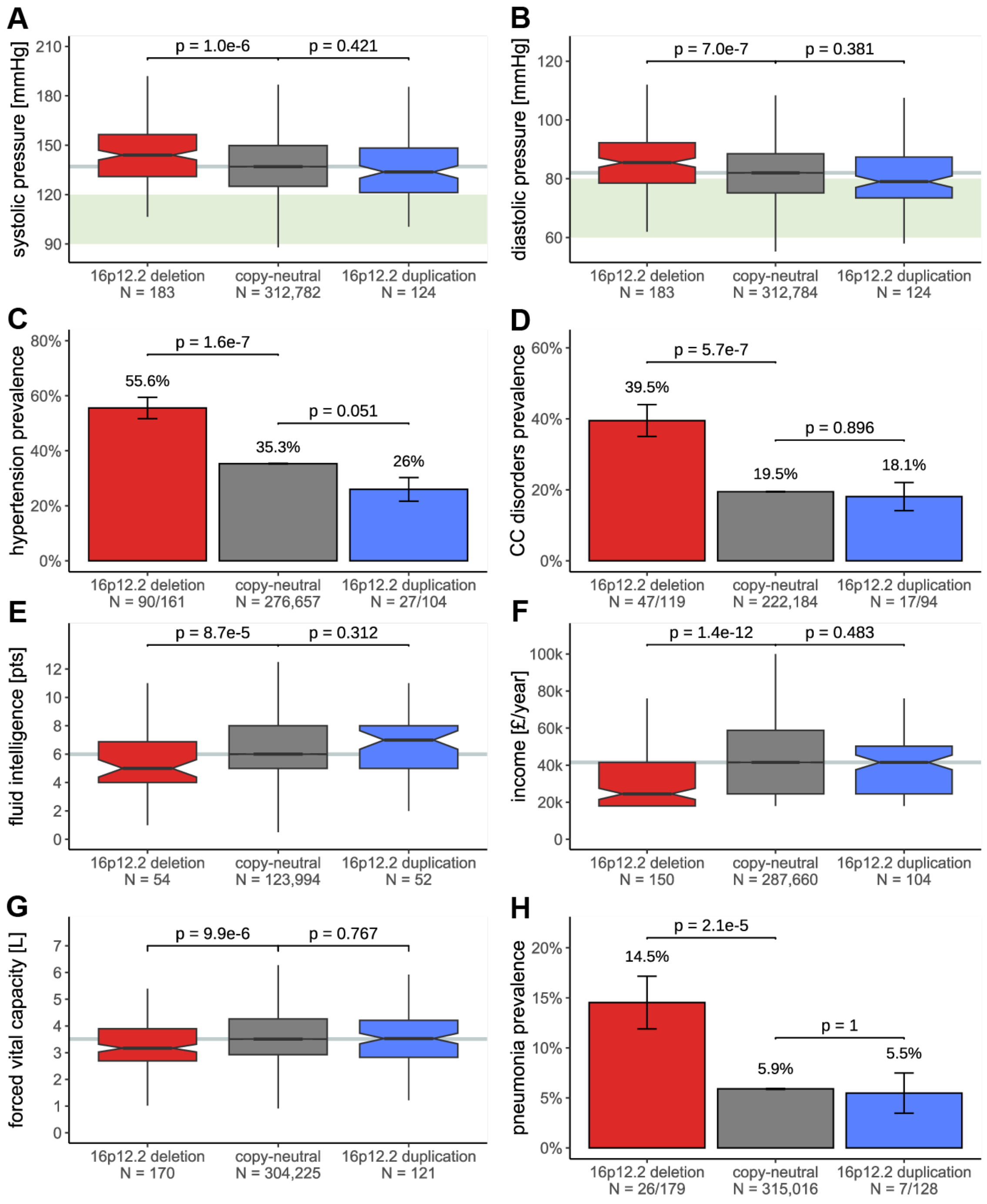


**Figure S4. 16p12.2 CNV region**

(**A**) Systolic and (**B**) diastolic blood pressure according to 16p12.2 copy-number (CN), shown as boxplots. Green background represents optimal blood pressure (systolic: 90-120 mmHg; diastolic: 60-80 mmHg). (**C**) Essential hypertension and (**D**) cardiac conduction (CC) disorders prevalence according to 16p12.2 CN, shown as bar plots. (**E**) Fluid intelligence score (maximum = 13 points), (**F**) average yearly total household income before taxes, and (**G**) forced vital capacity according to 16p12.2 CN, shown as boxplots. (**H**) Pneumonia prevalence according to 16p12.2 CN, shown as a bar plot. For boxplots, outliers are not shown; p-values compare deletion and duplication carriers to copy-neutral individuals (two-sided t-test); grey horizontal line represents median among copy-neutral individuals; sample sizes are indicated (N). For bar plots, error bars represent ± the standard error; p-values compare prevalence among deletion and duplication carriers to the one in copy-neutral individuals (two-sided fisher test); number of cases and sample sizes are indicated (N = cases/sample size).

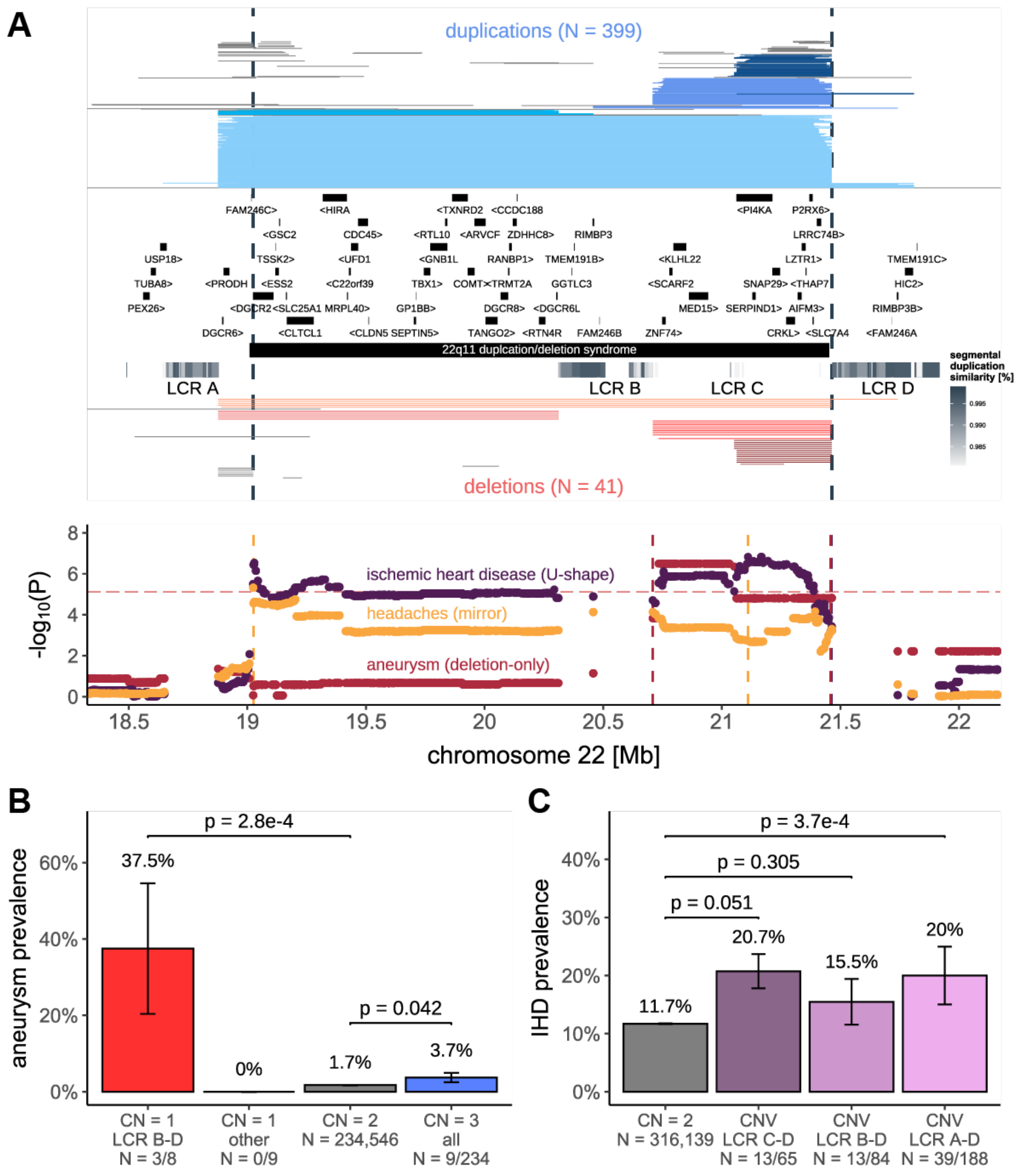


**Figure S5. 22q11.2 CNV region**

(**A**) 22q11.2 genetic landscape. Top: Coordinates of duplications (shades of blue; top) and deletions (shades of red; bottom) overlapping the maximal CNV region (CNVR delimited by vertical dashed lines) associated with ischemic heart disease (IHD), headaches, and aneurysm. CNVs are divided and colored according to 4 groups to reflect breakpoints at low-copy repeats (LCRs) spanning the region: A-D, A-B, B-D, C-D, with atypical CNVs in grey. LCRs are composed of segmental duplications, represented as a grey gradient proportional to the degree of similarity. Genomic coordinates of genes and DECIPHER CNV are displayed. Bottom: Negative logarithm of association p-values of CNVs (best model in parenthesis) with IHD, headaches, and aneurysm. Disease-specific CNVRs are shown with colored vertical dashed lines. Red horizontal dashed line represents the genome-wide threshold for significance for CNV-GWAS ($p \leq 7.5 \times 10^{-6}$). (**B**) Prevalence of aneurysm according to 22q11.2 copy-number (CN) and CNV group (A). P-values compare deletion (CN = 1) and duplication (CN = 3) carriers from various groups (other = A-D, A-B, C-D; all = A-D, A-B, B-D, C-D) to copy-neutral (CN = 2) individuals (two-sided Fisher test). (**C**) Prevalence of IHD according to CNV groups (A). P-values compare IHD prevalence among individuals carrying a CNV (duplication or deletion) spanning LCR C-D, B-D, or A-D to copy-neutral (CN = 2) individuals (two-sided Fisher test). (B, C) Error bars represent ± standard error; number of cases and sample sizes are indicated (N = cases/sample size).

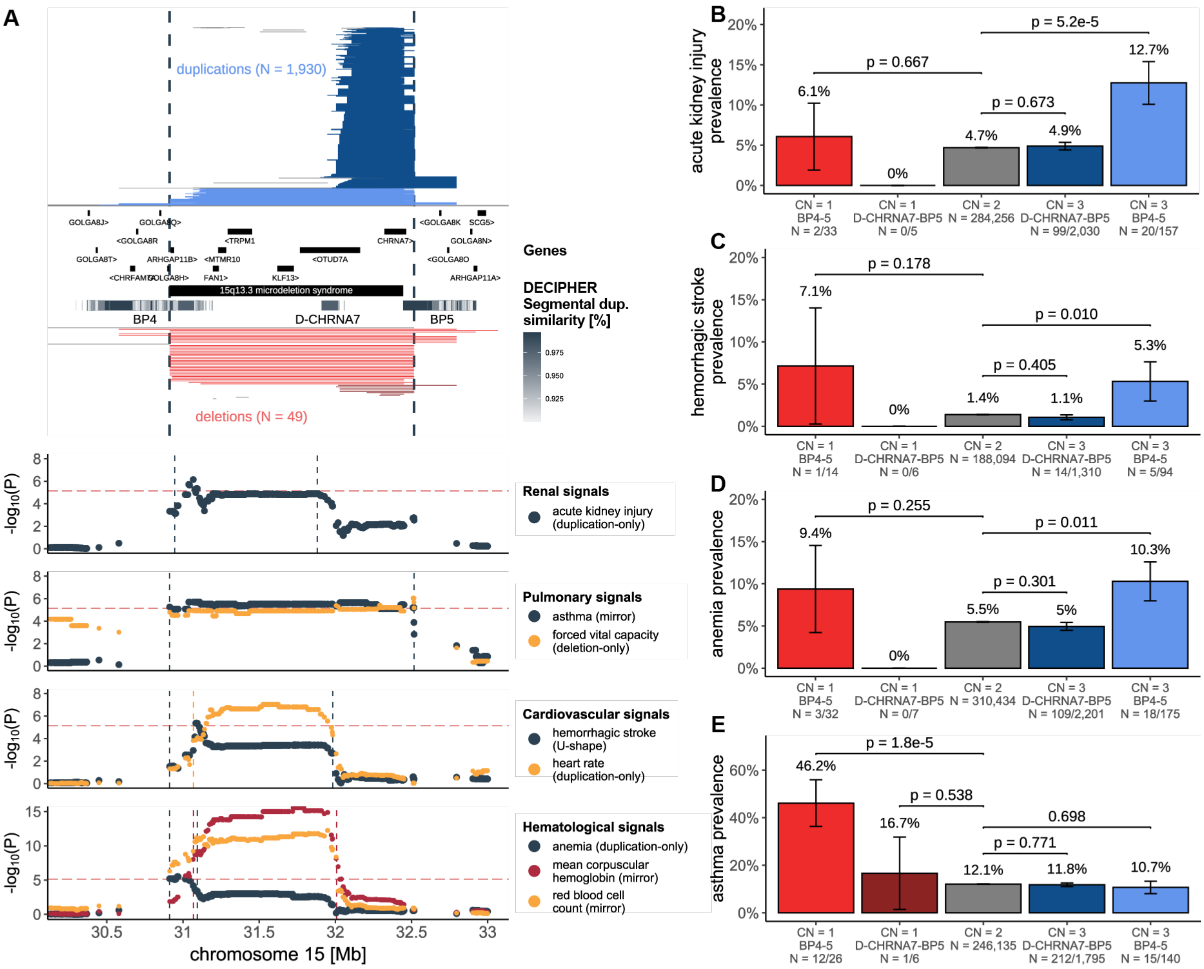


#### Figure S6. 15q13 CNV region

(**A**) 15q13 genetic landscape. Top: Coordinates of duplications (shades of blue; top) and deletions (shades of red; bottom) overlapping the maximal CNV region (CNVR delimited by vertical dashed lines) associated with acute kidney injury (AKI), asthma, forced vital capacity, hemorrhagic strokes, heart rate, anemia, mean corpuscular hemoglobin, and red blood cell count. CNVs are divided and colored according to whether they span breakpoint (BP) 4 to 5 or D-CHRNA7 to BP5, with atypical CNVs in grey. Breakpoints reflect segmental duplications, represented as a grey gradient proportional to the degree of similarity. Genomic coordinates of genes and DECIPHER CNV are displayed. Bottom: Negative logarithm of association p-values of CNVs (best model in parenthesis) with renal, pulmonary, cardiovascular, and hematological traits. Traits-specific CNVRs are shown with vertical dashed lines. Red horizontal dashed line represents the genome-wide threshold for significance for CNV-GWAS ($p \leq 7.5 \times 10^{-6}$). (**B**, **C**, **D**, **E**) Prevalence (± standard error) of (**B**) AKI, (**C**) hemorrhagic stroke, (**D**) anemia, and (**E**) asthma according to 15q13 copy-number (CN) and group (A). P-values compare BP4-5 and D-CHRNA7-BP5 deletion (CN = 1) and duplication (CN = 3) carriers to copy-neutral (CN = 2) individuals (two-sided Fisher test). Number of cases and sample sizes are indicated (N = cases/sample size).

### SUPPLEMENTAL NOTES

#### **NOTE 1:** Microarray-based CNV calling.

##### *CNV encoding in PLINK*

CNV matrices were encoded into three PLINK binary file sets (`--make-bed` PLINK v1.9; Supplementary Note 1 – Table 1) [1]. To reduce file size and facilitate parallelized computation, files are split at the chromosome level (i.e., for each PLINK encoding there are 24 files: 22 autosomes + pseudoautosomal regions + chromosome X). PLINK file sets were used to fit four association models mimicking different modes of CNV action: mirror, U-shape, duplication-only, or deletion-only (*Main CNV-GWAS model*).

| Association models | Mirror | | U-shape | | Duplication-only | | Deletion-only | |
|---|---|---|---|---|---|---|---|---|
| PLINK file set | $PLINK_{CNV}$ | | $PLINK_{CNV}$ | | $PLINK_{DUP}$ | | $PLINK_{DEL}$ | |
| Encoding | Num. | PLINK | Num. | PLINK* | Num. | PLINK | Num. | PLINK |
| Deletion (QS < -0.5) | -1 | AA | 1 | AA | NA | 00 | 1 | TT |
| Copy-neutral (\|QS\| ≤ 0.5) | 0 | AT | 0 | AT | 0 | AT | 0 | AT |
| Duplication (QS > 0.5) | 1 | TT | 1 | TT | 1 | TT | NA | 00 |

**Supplementary Note 1 – Table 1. Encoding of CNVs**
Encoding of high-confidence CNVs (|QS| > 0.5) into numerical probe-by-sample matrices (Num.) and three PLINK file set (PLINK), mimicking encoding of single-nucleotide variants. *The U-shape model uses the same PLINK file set as the mirror model but is assessed with the `hetonly` modifier of PLINK's `glm` function, allowing to compare the effect of deletion and duplications against copy-neutral individuals.

##### *Chromosome X CNVs*

Chromosome X CNVs were called using dedicated PennCNV modalities as previously described [1]. To avoid interference between the two-letter CNV encoding (Supplementary Note 1 – Table 1) and the male chromosome X hemizygosity assumption of PLINK, all individuals are (falsely) labeled as female when performing genetic analyses in PLINK.

#### **NOTE 2:** Extended phenotypic assessment.

For continuous traits, unless specified otherwise, average over available instances were used.

##### *BRCA1 deletion*

Medical history of female *BRCA1* deletion carriers is based on #41270 (*diagnosis – ICD10*) and age at diagnosis was calculated as previously described (*Case-control definition and age at disease onset calculation*). Relevant and prevalent diagnoses were manually selected for display. For the hereditary breast and ovarian cancer (HBOC) prevalence and time-to-event analysis, we considered C50 (malignant neoplasm of breast), C53 (malignant neoplasm of cervix uteri), C54 (malignant neoplasm of corpus uteri), C55 (malignant neoplasm of uterus, part unspecified), C56 (malignant neoplasm of ovary), and C57 (malignant neoplasm of other unspecified female genital organs) ICD-10 diagnoses on the inclusion list and used the same exclusion list as for ovarian cancer. Duplications and low-quality CNVs ($|QS| \leq 0.5$), as well as male individuals were excluded from the analyses. Difference in prevalence was assessed by two-sided Fisher test. Time-to-event analysis was performed as previously described (*Statistical confidence tiers*) to estimate the effect of the *BRCA1* deletion, using age, $age^2$, array, and PC1-40 as covariates.

##### *LDLR deletion*

Medical history of *LDLR* deletion carriers is based on #41270 (*diagnosis – ICD10*) and age at diagnosis was calculated as previously described (*Case-control definition and age at disease onset calculation*). Drug usage data originates from #20003 (*treatment/medication code*). The list of considered hypolipidemic agents and antihypertensive/antianginal drugs (Supplementary Note 2 – Table 1) was based on: https://www.drugs.com/ (29/09/2022). A minimum of 3 individuals was required for a code/drug to be displayed.

| Category | Description | UKBB_code |
|---|---|---|
| **statins** | atorvastatin | 1141146234 |
| | lipitor 10mg tablet | 1141146138 |
| | fluvastatin | 1140888594 |
| | lescol 20mg capsule | 1140864592 |
| | pravastatin | 1140888648 |
| | rosuvastatin | 1141192410 |
| | crestor 10mg tablet | 1141192414 |
| | simvastatin | 1140861958 |
| | zocor 10mg tablet | 1140881748 |
| | zocor heart-pro 10mg tablet | 1141200040 |
| | eptastatin | 1140910632 |
| | velastatin | 1140910654 |
| **cholesterol absorption inhibitors** | ezetimibe | 1141192736 |
| | ezetrol 10mg tablet | 1141192740 |
| **fibrates** | fenofibrate | 1140861954 |
| | gemfibrozil | 1140861856 |
| | gemfibrozil product | 1141157262 |
| | lopid 300 capsule | 1140861858 |
| | clofibrate | 1140861944 |
| | bezafibrate product | 1141157260 |
| | bezafibrate | 1140861924 |
| | bezalip 200mg tablet | 1140861926 |
| | bezalip-mono 400mg m/r tablet | 1140861928 |
| **bile acid sequestrants** | cholestyramine+aspartame 4g/sachet powder | 1140861942 |
| | cholestyramine | 1140865576 |
| | cholestyramine product | 1141157416 |
| | questran 4g/sachet powder | 1140861936 |
| | colestipol | 1140888590 |
| | colestid 5g/sachet granules | 1140861848 |
| **cardioselective beta-blocker** | atenolol | 1140866738 |
| | bisoprolol | 1140879760 |
| | cardicor 1.25mg tablet | 1141171152 |
| **ACE inhibitor** | ramipril | 1140860806 |
| | perindopril | 1140888560 |
| | lisinopril | 1140860696 |

**Supplementary Note 2 – Table 1. Considered drugs with UK Biobank encoding**
Drugs from #20003 considered in the displayed drug categories in Figure 4E. UK Biobank encoding is provided in the last column.

For prevalence and time-to-event analysis, only E78.0 (pure hypercholesterolemia) was considered on the inclusion list; the same exclusion list as for lipidemias was used. Duplications and low-quality CNVs (|QS| ≤ 0.5) were excluded from analyses. Difference in prevalence was assessed by two-sided Fisher test. Time-to-event analysis was performed as previously described (*Statistical confidence tiers*) to estimate the effect of the *LDLR* deletion, using sex, age, age$^2$, array, and PC1-40 as covariates.

Low-density lipoprotein (LDL) cholesterol measurements were available for seven LDLR deletion carriers in #42040 (*GP clinical event records*). P12 was excluded as the blood biochemistry LDL levels preluded the first primary care measurement. LDL levels of earliest measurement on record (primary care) was compared to LDL levels from standardized blood biochemistry measurement (#30780) taken at assessment (#53) using a one-sided paired t-test. Based on #42039 (*GP prescription records*), we were able to identify both P5 and P13 as being prescribed statins by their general practitioner despite no record of statin usage in #20003.

##### *17q12 deletion*

For time-to-event analysis, the same chronic kidney disease (CKD) definition as in the main analysis was used. Low-quality CNVs (|QS| ≤ 0.5) were excluded from analyses. Time-to-event analysis was performed as previously described (*Statistical confidence tiers*), modeling both 17q12 deletions and 17q12 duplications in the same CoxPH model, adjusted for sex, age, age$^2$, array, and PC1-40. Estimated glomerular filtration rate (eGFR) was calculated based on the CKD-EPI equation using #30700 (*creatinine* [μmol/L]), accounting for age, sex and ancestry [2].

##### *Other phenotypes*

Other used phenotypes include *alkaline phosphatase* (#30610), *systolic* (#4080) and *diastolic* (#4079) *blood pressure (automated reading)*, *forced vital capacity* (#3062), *fluid intelligence score* (#20016), and *average total household income before tax* (#738). The latter categorical variable was transformed to be continuous: ≤ £18,000 to £18,000; £18,000-30,999 to £24,500; £31,000-51,999 to £41,500; £52,000-100,000 to £76,000; ≥ £100,000 to £100,000.

### **NOTE 3:** CNV subgroup definition.

When analyzing complex CNVRs (i.e., 16p13.11, 22q11.2, 15q13), CNV carriers were split into subgroups based on visual inspection of their CNV breakpoints and segmental duplications overlapping the region. Criteria below were used to define groups (Supplementary Note 3 – Table 1). CNVs not matching any of the groups are referred to as "atypical" CNVs.

| CNVR | group | chr | min. start [bp] | max. start [bp] | min. end [bp] | max. end [bp] |
|---|---|---|---|---|---|---|
| **16p13.11** (Figure 5) | cat1 | 16 | - | 15,000,000 | 16,250,000 | 16,750,000 |
| | cat2 | 16 | 15,000,000 | 15,200,000 | 16,250,000 | 16,750,000 |
| | cat3 | 16 | 15,200,000 | 15,800,000 | 16,250,000 | 16,750,000 |
| | cat4 | 16 | - | 15,800,000 | 17,500,000 | - |
| | cat5* | 16 | 16,242,785 | - | - | 16,317,379 |
| **22q11.2** (Figure S5) | LCR A-D | 22 | 18,500,000 | 19,200,000 | 21,250,000 | 21,900,000 |
| | LCR A-B | 22 | 18,500,000 | 19,200,000 | 20,250,000 | 20,600,000 |
| | LCR B-D | 22 | 20,000,000 | 20,850,000 | 21,250,000 | 21,900,000 |
| | LCR C-D | 22 | 21,000,000 | 21,150,000 | 21,250,000 | 21,900,000 |
| **15q13** (Figure S6) | BP4-5 | 15 | 30,250,000 | 31,250,000 | 32,300,000 | 33,100,000 |
| | D-CHRNA7-BP5 | 15 | 31,700,000 | 32,300,000 | 32,300,000 | 33,100,000 |

**Supplementary Note 3 – Table 1. CNV carrier subgrouping**
Selection criteria for different CNV carrier subgroups considered for analyzed CNV regions (related Figure in parenthesis). Minimum and maximum start and end positions reflect the range in which the CNV breakpoints are to be for a given CNV to be assigned to a subgroup. "-" indicates open end. * For 16p13.11 cat5 CNVs, coordinates correspond to the coordinates of *ABCC6*. All positions are in hg19/GRCh37.

### SUPPLEMENTAL REFERENCES


1. Auwerx C, Lepamets M, Sadler MC, Patxot M, Stojanov M, Baud D, et al. The individual and global impact of copy-number variants on complex human traits. Am J Hum Genet. 2022;109:647–68.
2. Levey AS, Stevens LA, Schmid CH, Zhang Y, Castro AF, Feldman HI, et al. A New Equation to Estimate Glomerular Filtration Rate. Ann Intern Med. 2009;150:604.
